# Deep Learning-Based Screening for *POLE* mutations on Histopathology Slides in Endometrial Cancer

**DOI:** 10.64898/2026.02.06.26345335

**Authors:** Nikki van den Berg, Lydia Schoenpflug, Nanda Horeweg, Sarah Volinsky-Fremond, Jurriaan Barkey-Wolf, Sonali Andani, Maxime Lafarge, Gitte Ørtoft, Jan J. Jobsen, Rubina Razack, Kees Gerestein, Trudy Jonges, Cor D. de Kroon, Remi Nout, Dorine Tseng, Nienke Kuijsters, Melanie E. Powell, Pearly Khaw, Lois Shepherd, Alexandra Leary, Stephanie M. de Boer, Stefan Kommoss, Anne Sophie V.M. van den Heerik, Marie A.D. Haverkort, David Church, Marco de Bruyn, Vincent T. H. B. M. Smit, Ewout Steyerberg, Carien L. Creutzberg, Viktor H. Koelzer, Tjalling Bosse

## Abstract

*POLE* sequencing for somatic mutations (*POLE*mut) guides adjuvant therapy in endometrial cancer (EC), but cost and infrastructural considerations lead to limited uptake. Omission of *POLE* testing leads to unnecessary exposure to radiotherapy and/or chemotherapy. We developed POLARIX, a multiple instance deep learning model with attention pooling, which predicts *POLE* mutation status from routine hematoxylin and eosin whole-slide images (WSIs). Trained on 2,238 cases from eleven EC cohorts, POLARIX showed clinical-grade discrimination across three external cohorts (Pooled: AUC=0.95, 95% CI: 0.91–0.98; *n*=68/481 *POLE*mut/*POLE*wt). Attention maps highlight *POLE* morphologies. Clinical applicability is demonstrated using predefined thresholds based on three resource scenarios. The most sensitive threshold (“Low”) yields a test reduction of 77% (73%-81%) (sensitivity: 93% (85%-99%), specificity: 89% (87%-92%)). POLARIX is an interpretable and cost-efficient approach to reduce *POLE* testing in women with endometrial cancer, broadening access to precision oncology.

## Main text

Endometrial cancer (EC) is the most common gynecological malignancy worldwide, affecting 420,242 women in 2022, and is increasing in incidence^1,2^. It is typically managed by surgical resection, with postoperative adjuvant therapy guided by risk factors. Risk factors incorporated in current guidelines include FIGO stage, histological subtype, tumor grade, lymphovascular space invasion LVSI, and the EC molecular classification when available^3–5^. Evaluation of pathogenic exonuclease domain mutations in DNA polymerase epsilon (*POLE*) by DNA sequencing is an essential component of the EC molecular classification, as patients with *POLE* mutant (*POLE*mut) tumours have excellent prognosis and do not require adjuvant therapy^6–8^. Routine *POLE* testing is therefore recommended to ensure accurate stratification and prevent overtreatment.

Despite guideline recommendations, *POLE* testing is not routinely performed^9^. In high-resource settings, barriers to implementation include lack of reimbursement and long turnaround times^10^. In low-resource settings, costs, reagent shortages, and lack of laboratory infrastructure and capacity often restrict testing to a few specialized facilities^11^. Consequently, many thousands of women with EC worldwide are unable to access personalized treatment, and are exposed to unnecessary radiotherapy and/or chemotherapy^12^. Hence, there is a crucial need for accessible methods that can reduce the need for *POLE* DNA sequencing through preselection of patients requiring confirmatory testing.

Deep learning (DL) models can infer molecular alterations directly from routine hematoxylin & eosin (H&E)-stained slides^13–19^. DL models are cost-efficient and compatible with digital pathology workflows. Prediction of *POLE* mutation status in EC from H&E slides using DL has been explored in three prior studies, demonstrating proof-of-concept feasibility; however, distinction between *POLE*mut EC and MMR-deficient EC proved challenging and larger-scale studies with extensive external testing are essential before clinical implementation can be considered^20–22^.

We developed and validated POLARIX, a DL-based screening tool for *POLE*mut detection using routine H&E whole-slide images (WSIs) as input (Fig. 1). POLARIX requires pre-selection of MMR-proficient (MMRp) tumors, because this improves the discriminative performance to detect *POLE*mut tumors and the need for *POLE* sequencing is most pressing in patients with MMRp EC given the rarity of MMR-deficient (MMRd)-*POLE*mut tumors (<1%)^6,20,23,24^. POLARIX leverages a foundation model, H-optimus-1^25^, and uses an attention module to generate a continuous *POLE*mut probability^26^, the POLARIX-score. Scores are dichotomized to recommend either rule-out or perform confirmatory DNA sequencing by using one of three decision thresholds (Low, Mid, and High) balancing sensitivity and test reduction for three resource scenarios. In this way, POLARIX serves as a highly effective and fast pre-screening tool that selects only a small subset of EC patients for *POLE* DNA sequencing confirmation.

**Figure 1.**
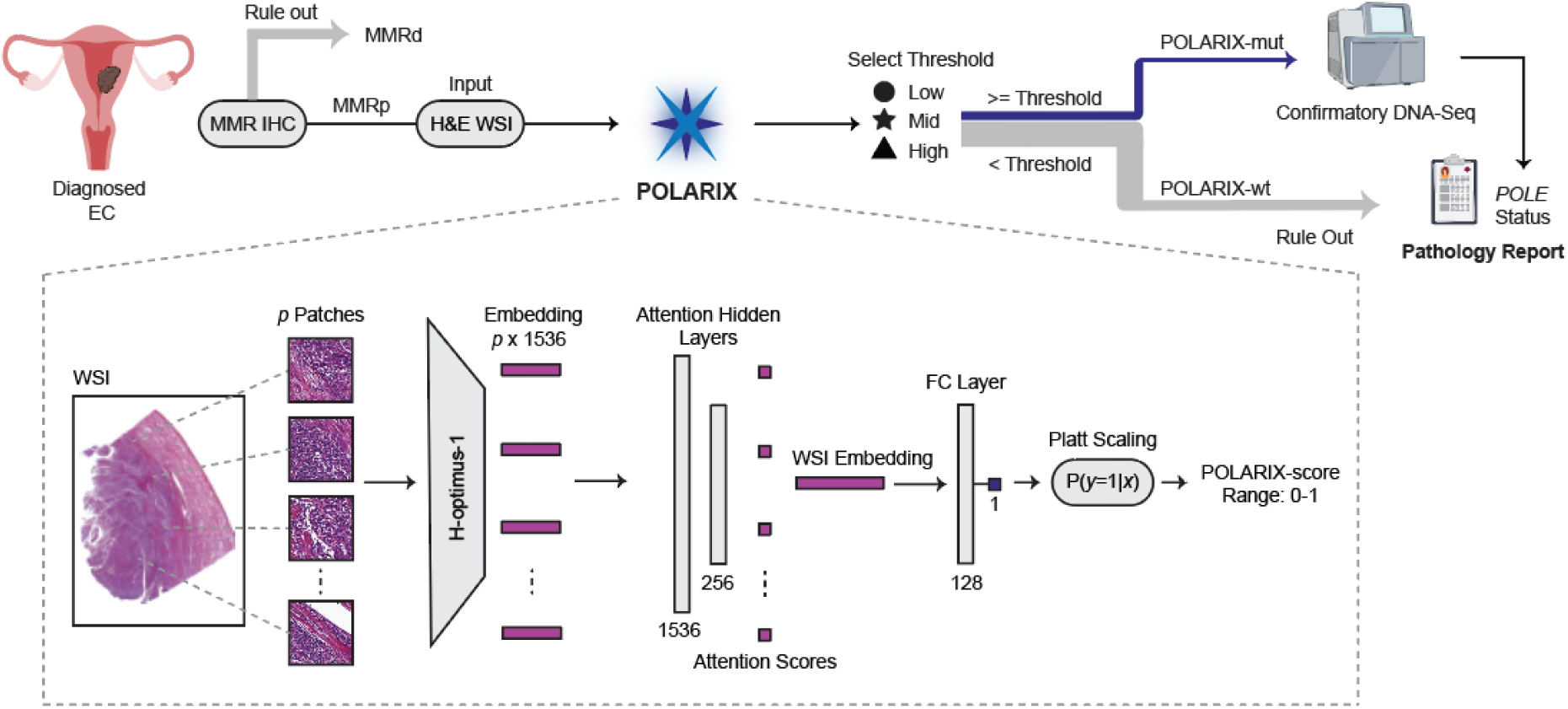
Clinical workflow and technical modules of POLARIX. First, MMR-proficient (MMRp) cases are selected with MMR immunohistochemistry (IHC) (MMRd = MMR-deficient). After selection, a digitized hematoxylin & eosin (H&E) whole-slide image (WSI) is used as input for POLARIX to extract 1536-dimensional H-optimus-1^25^ embeddings from 180 µm patches at 40× magnification (0.25 µm/px). Then, a multiple instance learning network with attention pooling predicts the likelihood of *POLE*mut, the POLARIX-score. Patch embeddings are weighted by a gated attention and aggregated into a slide-level representation^26^, followed by a fully-connected (FC) layer and a sigmoid classifier producing a continuous probability between 0 and 1. This continuous prediction is then dichotomized using one of the decision thresholds (Low, Mid or High), and allocates patients as POLARIX-wildtype (POLARIX-wt) requiring no further testing or as POLARIX-mut that requires confirmatory *POLE* DNA sequencing.

## Results

### Cohorts

We assembled the largest and most diverse EC dataset worldwide, comprising 4,187 patients with known *POLE* status (Supplementary Fig. 1). Among these, 3,010 EC were MMRp, and 2,719 had representative hysterectomy WSIs suitable for analysis. Model development used data from 2,238 patients from three randomized trials^27–29^, six retrospective cohorts^30–35^, and two public datasets^36–38^, capturing demographic, technical, and biological variation (Extended Fig. 1).

External testing was performed on data from 481 patients from three cohorts, the randomized high-risk trial PORTEC-3^39^, the prospective population cohort from Medisch Spectrum Twente (MST-II), and the retrospective high-risk cohort from University Medical Center Utrecht (UMCU-HR)^40^ (Extended Fig. 1). We pooled the PORTEC-3, MST-II, and UMCU-HR cohorts into a single “Pooled” external test set, and reported performances on cohorts separately. This design enabled rigorous evaluation across trials, institutions, and patient risk groups, reflecting the variability expected in real-world deployment.

**Extended Figure 1.**
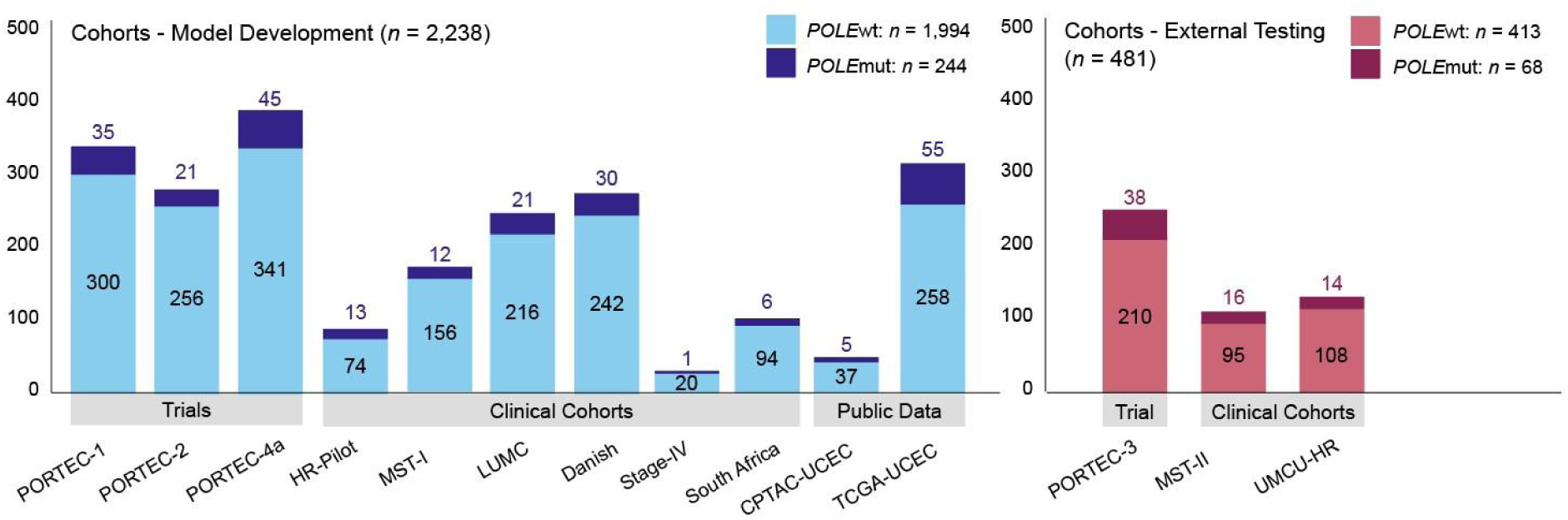
We included one representative WSI per patient from a formalin-fixed, paraffin-embedded (FFPE) block. The model development dataset (*n* = 2,238 patients total, 244 *POLE*mut) consisted of three randomized trials, six clinical cohorts, and two public datasets. The external test cohorts (*n* = 481 patients total, 68 *POLE*mut) consisted of PORTEC-3, MST-II, and UMCU-HR.

### Performance of POLARIX

Testing on the pooled external cohorts (n = 481) showed an area under the curve (AUC) of 0.95 (95% CI: 0.91–0.98) (Fig. 2a), indicating excellent discrimination between *POLE*mut and *POLE*wt cases. Moreover, performance was consistently high across each of the external testing cohorts, with AUCs of 0.97 (0.95–0.99) in PORTEC-3, 0.90 (0.75–0.99) in MST-II, and 0.96 (0.93–0.99) in UMCU-HR (Fig. 2b-d).

**Figure 2.**
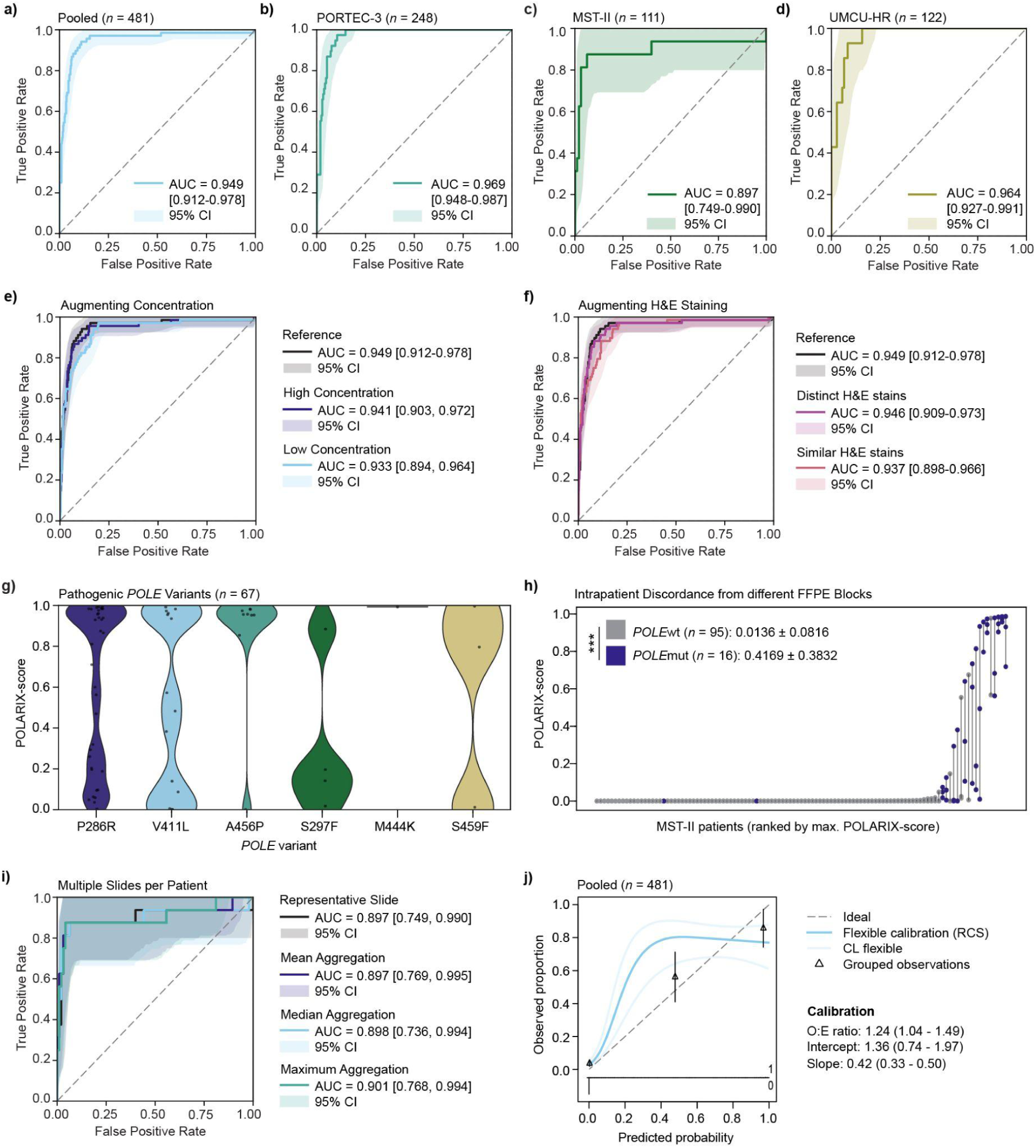
Performance of POLARIX in external testing. **a-d)** Discriminative performance quantified by the AUC. **e-f)** Performance of POLARIX under simulated H&E stain concentration and colour variation. **g)** POLARIX scores per pathogenic *POLE* variant in the pooled external test cohort. Descriptives in Supplementary Table 4 and comparisons between distributions in Supplementary Table 5. **h)** Intrapatient discordance in the MST-II cohort. POLARIX-scores were plotted from slides originating from different FFPE blocks, with the discordance represented as the connecting line for each patient. Blue dots represent true *POLE*mut patients, grey *POLE*wt. **i)** Discriminative performance for the multiple slide per patient scenario, using the MST-II slides from different FFPE blocks. **j)** Calibration plot depicting the ideal calibration and POLARIX calibration, approximated with flexible calibration (Methods: Statistical Analysis).

To evaluate prediction robustness, we first evaluated robustness to staining variability (Supplementary Fig. 3-4). Performance declined slightly under conditions with low concentration and high colour similarity, but showed robustness to staining variability across all settings (AUC > 0.93) (Fig. 2e-f). Thereafter, we performed a *POLE* variant analysis, which showed that non-pathogenic *POLE* variants (*n* = 3) yielded POLARIX-scores near zero, and indicated that POLARIX score distributions did not significantly differ between pathogenic *POLE*mut variants (Fig. 2g and Supplementary Table 4-5). Next, we tested the intra-patient concordance in the MST-II cohort, for which we had two (*n* = 9) or three (*n* = 102) WSIs from different FFPE blocks per patient available. Here, we found that intrapatient concordance was high in the true *POLE*wt cases (POLARIX-score = 0.014 ± 0.082), which is important as POLARIX-wt cases would not get confirmatory DNA sequencing in our design (Fig. 2h). There was more variability in the true *POLE*mut cases (POLARIX-score = 0.42 ± 0.38), but on average they yield significantly higher POLARIX-scores than true *POLE*wt cases. Lastly, we leveraged the MST-II cohort to also investigate the best approach for the selection of slides. We found that selecting the slide with the highest POLARIX-score yielded the highest AUC (0.901, CI: 0.768-0.994), but this was only marginally higher than using a representative slide (AUC: 0.897 (0.749-0.990) (Fig. 2i).

Next, we evaluated the calibration of POLARIX to determine whether the predicted POLARIX-scores correspond to the prevalence of true *POLE*mut cases. We first confirmed that Platt-scaling improved calibration during model development (Supplementary Table 6 and Supplementary Figure 5). On the external pooled cohort, the model slightly underestimated risk on average, with an O:E ratio (ideal = 1) of 1.24 (CI: 1.04-1.49) (Fig. 2j). The intercept (ideal = 0) was 1.36 (0.74-1.97) and the slope (ideal = 1) was 0.42 (0.33-0.50). Grouped-observation calibration showed agreement between predicted and observed probabilities in the lower-to-intermediate risk range: low-risk (predicted 0.4%, observed 1.6% (CI: 0.7-3.4)), intermediate-risk (predicted 48%, observed 47% (34-60)), and high-risk (predicted 98%, observed 77% (64-87)). Separate calibration assessments on external test cohorts (PORTEC-3, MST-II and UMCU-HR) are depicted in (Supplementary Table 7) and (Supplementary Figure 6).

### Determining and evaluating decision thresholds

POLARIX was evaluated using three predefined decision thresholds for three resource scenarios: maximizing sensitivity for safe rule-out (threshold Low), a balance between sensitivity and DNA sequencing rule-out (threshold Mid) and prioritizing DNA sequencing test reduction with acceptable sensitivity (threshold High). Thresholds were defined based on clinically acceptable trade-offs between true-positive and false-positive classifications. Threshold Low was set at a POLARIX-score of 0.005, corresponding to acceptance of 200 false-positive cases (unnecessary DNA sequencing of *POLE*wt tumours) to identify one true-positive *POLE*mut tumour. Threshold Mid was set at 0.025, corresponding to acceptance of 40 false positives per true positive, and threshold High at 0.075, corresponding to acceptance of 13 false positives per true positive.

POLARIX-scores were dichotomized across a range of classification thresholds (0.00–0.10), and for each decision threshold we quantified missed *POLE*mut cases (false negatives) and DNA sequencing test reduction (true negatives). Across all thresholds, POLARIX demonstrated strong classification performance in the pooled external test set (Fig. 3a–b). At threshold Low, test reduction was reduced by 77%, with 9.1% of cases classified as false positives (i.e., unnecessarily DNA sequenced *POLE*wt tumours). Five of 68 *POLE*mut cases were missed, corresponding to a sensitivity of 0.93 (0.85–0.99). Mid yielded a test reduction of 80% and had 6.4% false positives. The sensitivity was 0.88 (0.80–0.95) (8/68 missed *POLE*mut cases). Threshold High achieved 81% test reduction with 4.8% false positives. The sensitivity was 0.83 (0.73–0.91) (12/68 missed *POLE*mut cases). Results for each test cohort were consistent (Supplementary Table 8).

**Figure 3.**
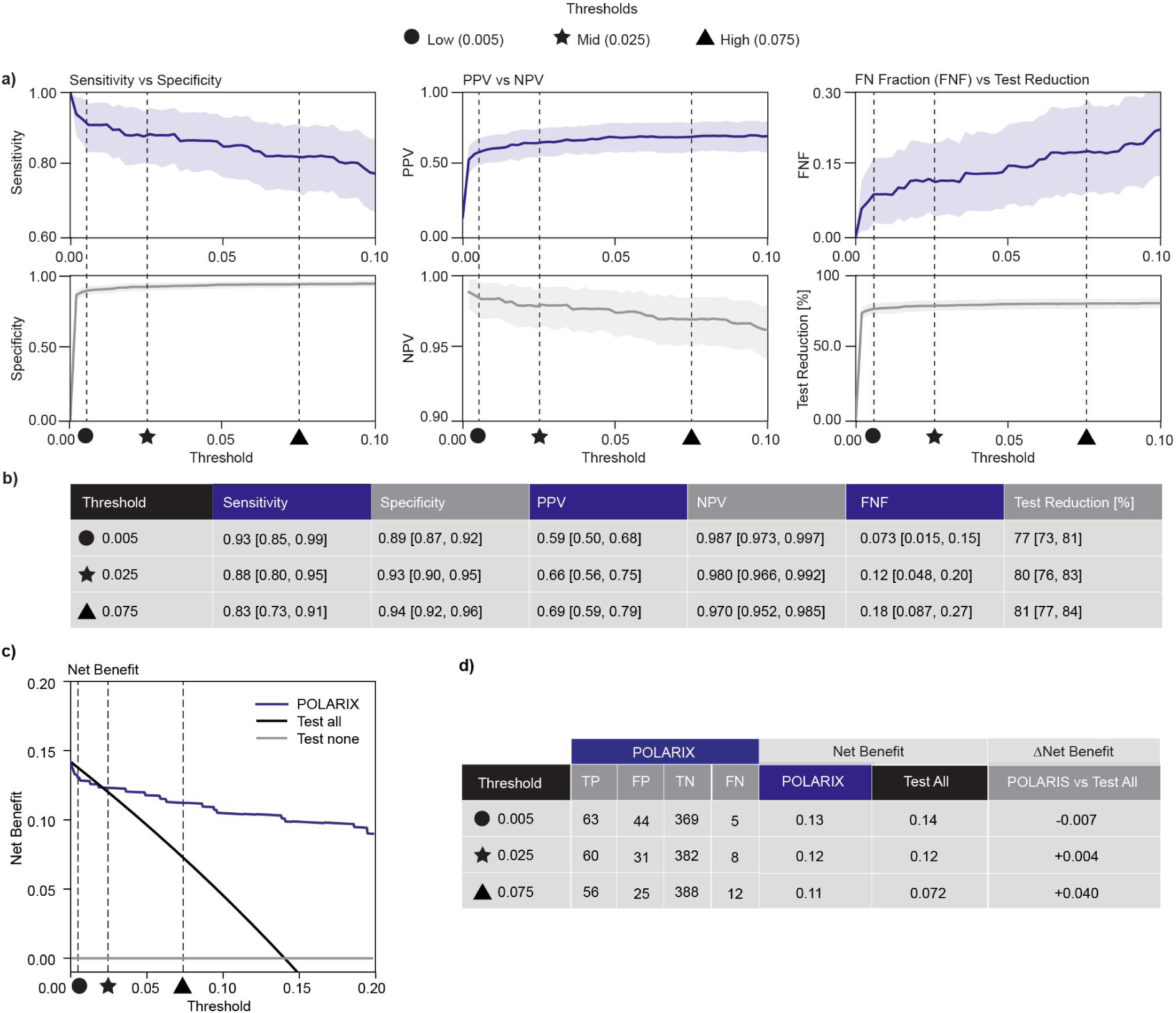
Classification performance and net benefit computation. **a)** Classification performance was evaluated by dichotomizing POLARIX-scores across a continuous threshold range (0.00–0.10) into POLARIX-mutant and POLARIX-wildtype groups. Reported metrics include sensitivity, specificity, positive predictive value (PPV), negative predictive value (NPV), test reduction in percentages by calculating the true-negative (TN) fraction, and false-negative fraction (FNF), which is the proportion of missed true *POLE*mut cases. **b)** Classification metrics with 95% confidence intervals at thresholds Low, Mid, and High. **c)** Decision curve analysis (DCA) was used to evaluate the clinical utility of POLARIX compared to “test all” and “test none”. The “Net Benefit” is plotted, where a positive net benefit indicates a net gain in correctly identified cases, while a negative net benefit suggests more harm than benefit. **d)** Net benefit was calculated at the thresholds Low (0.005), Mid (0.025), and High (0.075), and compared to the “test all” and “test none” strategies. In this comparison, ΔNet Benefit is positive if the model is more beneficial than “test all” or “test none”. TP = true positives, FP = false positive, TN = true negative, FN = false negative

Clinical utility was assessed with decision curve analysis^41^ (DCA) (Fig. 3c-d), which compares POLARIX-based screening to two reference scenarios: DNA sequencing all patients (“test-all”) or none (“test-none”). Net benefit quantifies how much a testing strategy helps clinicians and patients by weighing the benefit of correctly identifying *POLE*mut cases against the cost of unnecessary sequencing in *POLE*wt cases. Hence, a higher net benefit means that the strategy achieves more clinical value for the same or fewer tests. Across Low, Mid, and High decision thresholds, POLARIX consistently provided greater net benefit than the “test-none” scenario, and outperformed the “test-all” scenario at the Mid and High thresholds. At Low, POLARIX performed slightly lower to “test-all” (ΔNet Benefit = −0.01), while at Mid and High it maintained strong net benefit (Net Benefit = 0.12 and 0.11, respectively) as the value of testing all patients declined. DCA results for each test cohort were consistent (Supplementary Table 9) and (Supplementary Fig. 7).

### Clinicopathological and morphological correlates of POLARIX-scores

To investigate which patient and tumour characteristics and morphological features are associated with POLARIX-scores, we leveraged the high-quality metadata of the PORTEC-3 trial, one of our three external test cohorts. This high-risk trial included stage II-III or stage I tumours with risk factors such as high tumor grade, non-endometrioid histotype and/or lymphovascular space invasion (LVSI) (Supplementary Table 2). Among the reference group (FIGO Stage I, Grade 1, endometrioid histotype (EEC) tumours, p53-wildtype, no LVSI, negative L1 cell adhesion molecule (L1CAM), and positive estrogen- (ER) and progesterone receptor (PR), the expected POLARIX score was 0.20 (95% CI: 0.08–0.33). After adjusting for all variables and relative to their respective reference categories, higher POLARIX-scores correlated with Grade 3, and lower POLARIX-scores correlated with p53-abnormality, FIGO Stage II and III, and clear cell carcinoma histotype (Fig. 4a–b, Supplementary Table 10).

**Figure 4.**
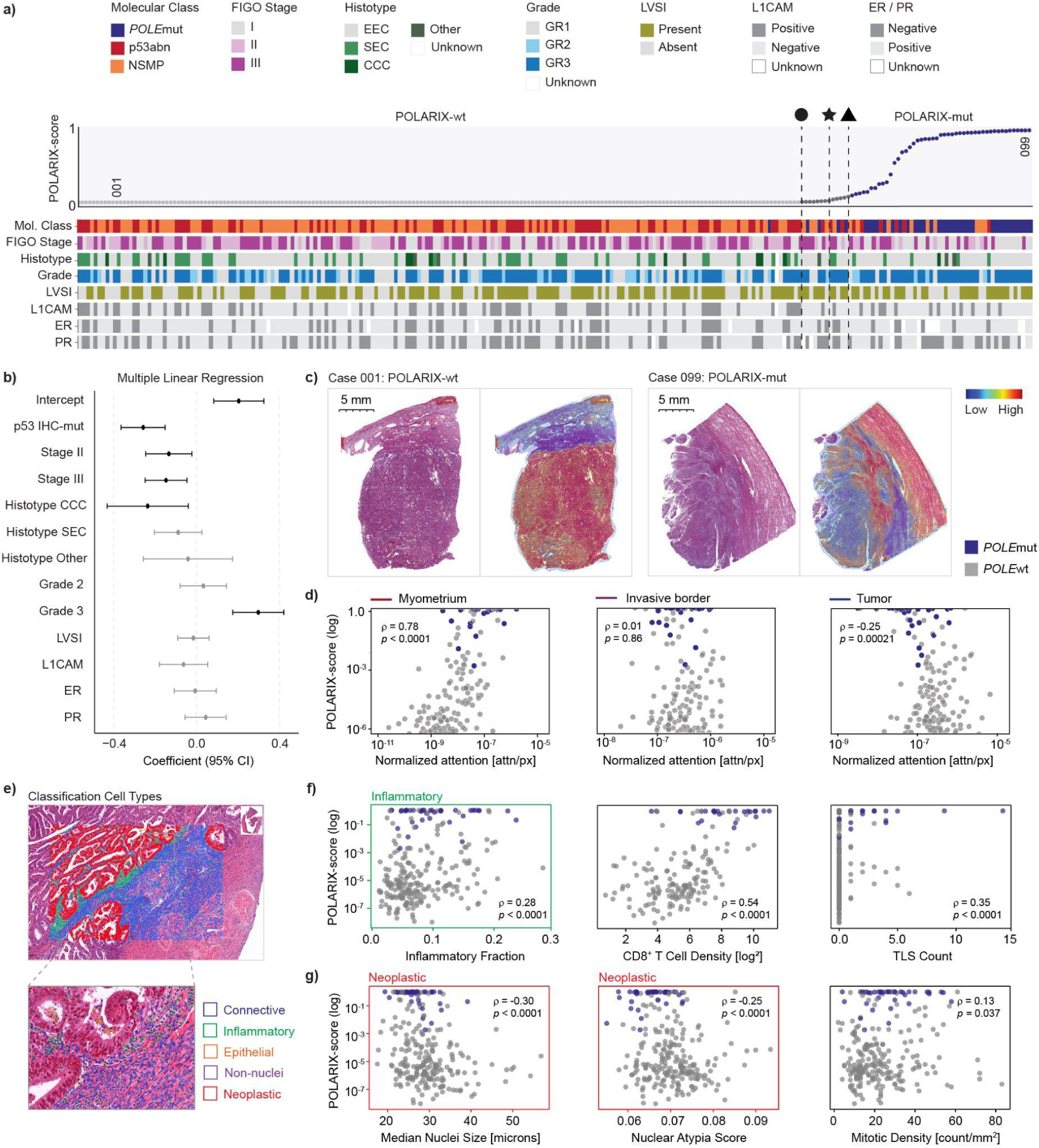
Clinicopathological and morphological correlates of POLARIX-scores. **a)** Heatmap of associations between clinicopathological covariates and continuous POLARIX-scores. **b)** Multiple linear regression analysis of clinicopathological variables and POLARIX scores. Coefficients represent adjusted mean differences in POLARIX score relative to reference groups: Stage I, Grade 1, and endometrioid histotype (EEC). Error bars represent 95% confidence intervals. **c)** Representative examples of attention heatmaps for POLARIX-wt and POLARIX-mut cases. **d)** Normalized attention distributions across annotated regions (tumor, invasive border, myometrium) (*n* = 248 WSIs). **e)** Representative example of a CellViT^++^ cell classification result. **f)** Correlations between POLARIX-scores and inflammatory features, including inflammatory cell fraction (*n* = 248 WSIs), CD8+ T-cell density (n = 168 TMAs), and tertiary lymphoid structure (TLS) counts (*n* = 238 WSIs). **g)** Correlations between POLARIX-scores and tumor nuclear features, including median nuclear area (*n* = 248 WSIs), nuclear atypia score (average of non-convexity and non-sphericity) (*n* = 248 WSIs), and mitotic density (*n* = 247 WSIs). Abbreviations: NSMP: no specific molecular profile, EEC: endometrioid histotype, SEC: serous endometrial carcinoma, CCC: clear cell carcinoma, GR: grade, LVSI: lymphovascular space invasion, L1CAM: L1 cell adhesion molecule, ER: estrogen receptor, PR: progesterone receptor, TLS: tertiary lymphoid structure.

We analyzed attention maps to identify whether specific slide regions drive predictions. The attention per annotated tumor, invasive border, and myometrial regions was quantified (Supplemental Fig. 8), which showed that POLARIX-scores correlated positively with myometrial attention, did not correlate with attention to the invasive border, and correlated negatively with tumor attention (Fig. 4d). Examples of representative cases are provided in (Fig. 4c).

Next, we examined immune-related features and tumor nuclear characteristics using available metadata or computed using CellViT^++^ (Supplemental Fig. 9). POLARIX-scores showed strong positive correlations with the fraction of inflammatory cells in whole-slide images (WSIs) (Fig. 4e), CD8+ T-cell density, and tertiary lymphoid structure (TLS) counts (Fig. 4f). Conversely, tumor nuclear characteristics reflecting atypia, including larger nuclear area, higher atypia scores, and higher mitotic density, were inversely correlated with POLARIX-scores (Fig. 4e, g).

### Misclassification analysis

To investigate misclassifications by POLARIX, we compared the morphological and immune features with a significant relation with the POLARIX score across true positives (TP), true negatives (TN), false positives (FP), and false negatives (FN) using ground-truth *POLE*mut status (Supplementary Tables 11-12).

*POLE-*wildtype cases with a false positive call by POLARIX had relatively dense intratumoral CD8^+^ cell infiltrates; significantly higher than the *POLE* wildtype cases with true negative call and comparable to those of true positive cases (Extended Figure 2a). Among tumor cell-related features, median nuclear area was comparable between *POLE* wildtype cases with a false positive call and true positive cases and lower than in *POLE* wildtype cases with true negative call (Extended Fig. 2a). Although inflammatory cell fraction, mitotic density, and nuclear atypia differed between POLARIX-mut and POLARIX-wt overall, *POLE* wildtype cases with a false positive call and true positive cases did not differ significantly for these features. Extended Fig. 2b shows a representative *POLE* wildtype case with a false positive call with TLS at the invasive border and within the myometrium, increased inflammatory cell density, small nuclear size, low nuclear atypia, and elevated mitotic activity, consistent with patterns observed in Fig. 4.

*POLE* mutant cases with a false negative call by POLARIX occurred at the Low, Mid, and High thresholds *n* = 1/3/5 times out of 38 true *POLE* mutant cases, respectively. These small numbers precluded group-level comparisons. Instead, all five cases were examined in detail (Supplementary Figure 10). We could not find general patterns describing these cases. One case showed lack of myometrium and a faint slide (Extended Fig. 2c).

**Extended Figure 2.**
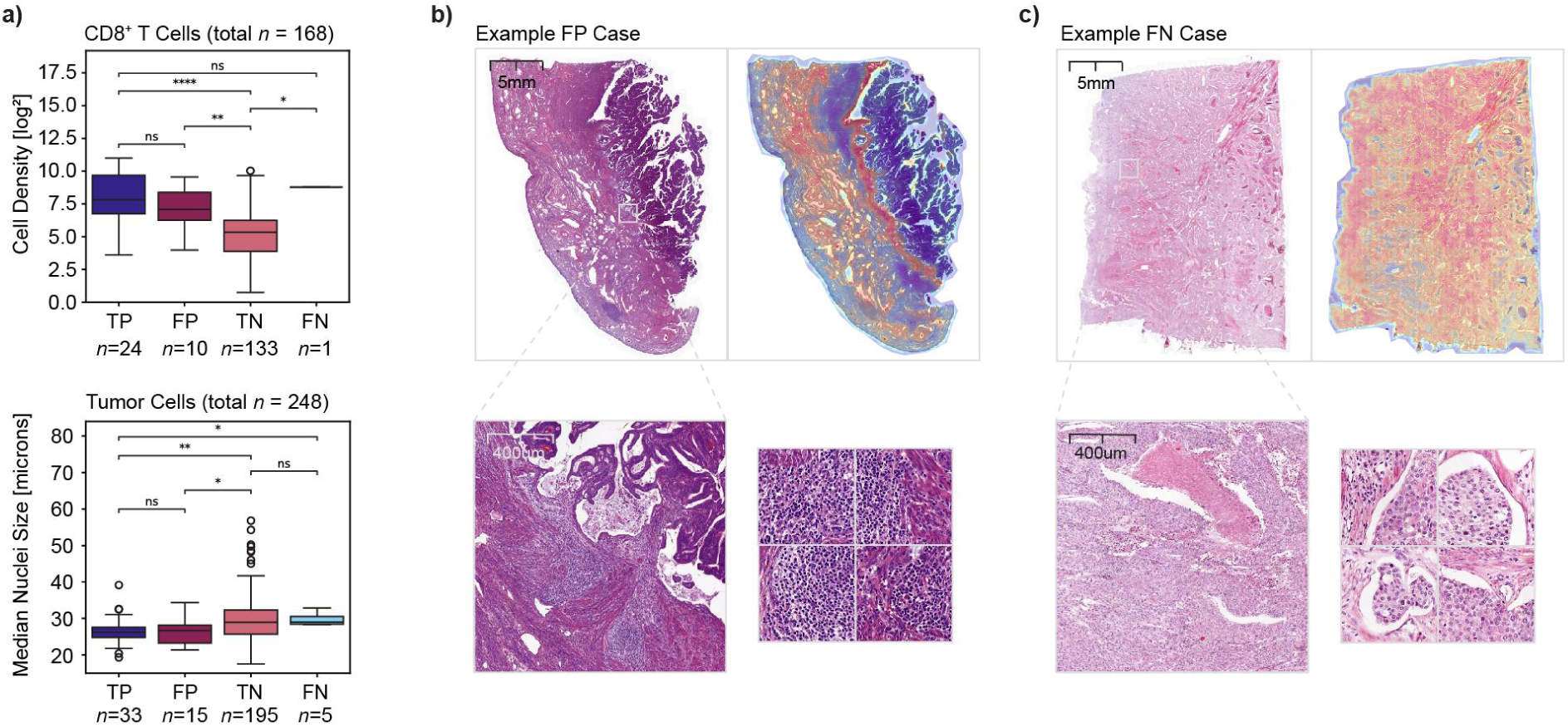
Characterizing misclassifications of POLARIX. **a)** Distributions of CD8+ T cell density and tumor cell nuclei size in prediction groups in PORTEC-3 (applying threshold High). **b)** Representative case of a false-positive call in *POLE*wt. **c)** Representative case of a false-negative call in *POLE*mut.

### Simulation of the implementation of POLARIX using PORTEC-3 data

To assess the clinical impact of POLARIX, we performed a simulation of deployment in PORTEC-3, an international trial including high-risk EC. We included 298 patients with clinical outcome data, from which 248 had MMRp tumours determined by IHC (Fig. 5a). POLARIX scores were dichotomized using the predefined Low, Mid, and High thresholds which subsequently lead to recommendations for confirmatory DNA sequencing or no further testing. Using known DNA sequencing results, we quantified benefits (*POLE*mut cases correctly identified for potential treatment de-escalation) and harms (*POLE*mut cases misclassified as POLARIX-wt, who might receive unnecessary radiotherapy or chemotherapy).

**Figure 5.**
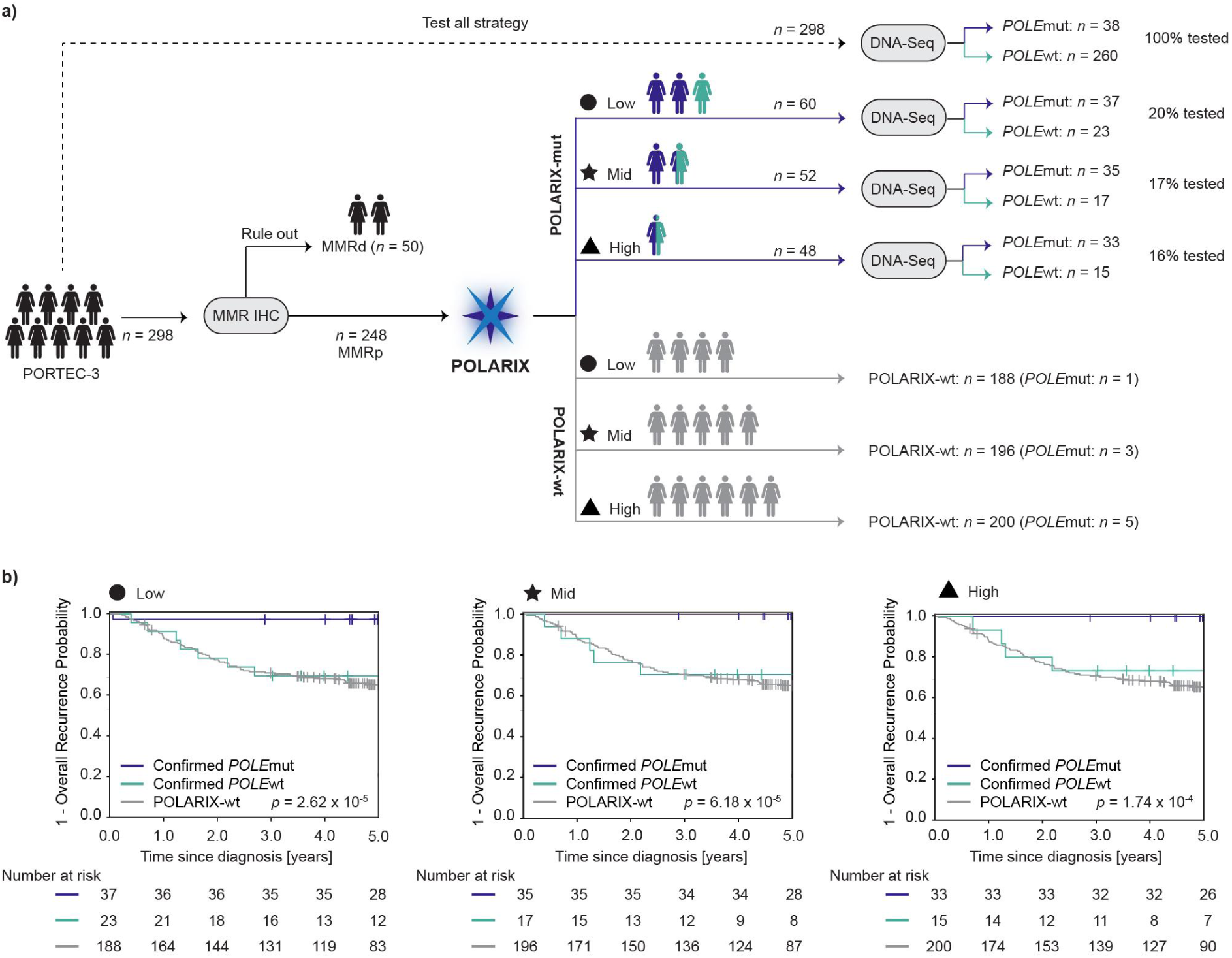
Simulated implementation of POLARIX in PORTEC-3. **a)** Patient flow when using the test all strategy, or use POLARIX with Low/Mid/High threshold, depicted with number tested of total. **b)** Five-year overall recurrence-free Kaplan Meier estimates stratified by the three possible scenarios: 1) POLARIX-mut followed by DNA sequencing *POLE*mut, 2) POLARIX-mut followed by DNA sequencing *POLE*wt, and 3) POLARIX-wt and no further DNA sequencing testing. Colours women icons: black for before running POLARIX, navy for confirmed *POLE*mut, green for confirmed *POLE*wt, grey for POLARIX-wt.

POLARIX reduced DNA sequencing use by 80% (threshold Low), 83% (threshold Mid) or 84% (threshold High), while preserving the prognostic stratification achieved by DNA-confirmed *POLE* testing (Fig. 5a). The Low threshold required confirmatory sequencing in 20% of patients and yielded potential de-escalation benefits in 37 patients against one patient potentially harmed due to a false-negative result (sensitivity 37/38, 97%). The Mid threshold required sequencing in 17% of patients, benefiting 35 and harming three. The High threshold required sequencing in 16% of patients, benefiting 33 and harming five. Importantly, the 5-year recurrence-free estimate for POLARIX-identified *POLE*mut EC with subsequent confirmation of *POLE*mut by sequencing remained high across thresholds (Low 97% (95% CI: 82–100), Mid 100% (100–100), High 100% (100–100)) (Figure 5b and Supplementary Table 13).

Kaplan-Meier analyses showed lower recurrence rates for POLARIX-mut vs POLARIX-wt patients, especially when mutation status was aligned with DNA sequencing results (Supplementary Figs. 11-12).

## Discussion

While EC incidence continues to rise worldwide^1,2^, the subgroup of patients with *POLE*mut EC with an excellent prognosis often remains unidentified due to high DNA sequencing costs, limited reimbursement, reagent shortages and restricted laboratory capacity^10,11^. Consequently, many women receive unnecessary radiotherapy or chemotherapy each year^7^. To prevent this, we developed POLARIX, a deep learning model that predicts *POLE* mutation status directly from routine H&E slides. With high sensitivity, high negative predictive value, and three resource-specific decision thresholds, POLARIX safely selects patients for confirmatory DNA sequencing while significantly reducing test demands.

Across external test cohorts, POLARIX demonstrated robust discriminative performance, with AUCs of 0.95 (95% CI, 0.91–0.98) in the pooled cohorts, 0.97 (0.95–0.99) in PORTEC-3, 0.90 (0.75–0.99) in MST-II, and 0.96 (0.93–0.99) in UMCU-HR. Prior developed DL models for *POLE* detection showed limited generalization (AUC was 0.68 (0.42–0.94); evaluated on ≈5 *POLE*mut cases^21^), limited assessment of robustness (AUCs were 0.97 and 0.98, no CI; 3/53 and 13/83 *POLE*mut test cases^22^), or lower performance, likely due to the complexity of directly predicting all EC molecular subtypes (AUC was 0.85; 51 *POLE*mut test cases)^20^. We hypothesize that the performance of POLARIX surpasses all prior DL models for *POLE* detection because of the large number of *POLE*mut cases we had available for training, the use of a foundation model (H-Optimus-1^25^) for feature extraction, and the pre-selection of only mismatch-repair proficient tumours. POLARIX further demonstrates robustness to staining variability and consistent score distributions across *POLE*mut variants. There was also high intrapatient concordance across multiple slides from different FFPE blocks in *POLE*wt cases, which is critical as POLARIX-wt cases would not get confirmatory DNA sequencing. Calibration assessment of POLARIX showed slight underestimation on average, and grouped-observation calibration showed moderate agreement between predicted and observed probabilities, both likely due to the rarity of the *POLE*mut class. This calibration assessment is necessary and omitted in the three previously discussed *POLE*mut prediction studies in EC^20–22^. Other proposed DL-based molecular alteration prediction models also did not report calibration^13,14,16–19^, except for the validation study of DL-based microsatellite instability prediction in colorectal cancer^42^. These findings establish POLARIX as the first strong candidate for a clinically robust screening solution for *POLE*mut detection.

Explainability analyses confirmed that POLARIX captures the immune-infiltrated phenotype and characteristic tumor morphology of *POLE*mut EC. At the cellular level, POLARIX-scores correlated positively with TLS counts, CD8^+^ T-cell density, and inflammatory cell fraction, consistent with previously reported associations with *POLE*mut tumors^43,44^. Attention mapping revealed strong correlations with myometrial regions, potentially reflecting immune-related microenvironmental features. In contrast, lower POLARIX-scores correlated with increased model attention toward tumor regions, nuclear atypia, and higher mitotic density, all features characteristic of aggressive EC^45–47^. The confirmation that POLARIX captures known morpho-molecular correlates enhances transparency and trust, which is important for POLARIX’ acceptability.

POLARIX was developed with three pre-defined resource-specific decision thresholds (Low, Mid, High), each demonstrating strong overall performance. At this moment, POLARIX-scores should be used in conjunction with these decision thresholds and not be interpreted as absolute probabilities of *POLE*mut. Across thresholds, sensitivity in the pooled cohort ranged from 83-93%, and DNA sequencing was reduced by 77-81%, even without considering the MMR-deficient cases that do not require POLARIX nor sequencing in our design. If the MMRd EC would be considered, the test reduction further increases to 80-84%. These test reductions outperform other DL-based pathology screening tools with decision thresholds, such as MSI detection in CRC that achieved a test reduction of almost half (specificity was 0.47, sensitivity was 0.97)^42^, and EGFR mutation screening in lung carcinoma that achieved a test reduction of 43% (PPV was 98%)^13^.

Among the three thresholds, we recommend using the Low threshold, which achieves clinically meaningful sensitivity of 93% (85-99), reduces confirmatory testing by 80% (including pre-selection of MMRp patients), and preserves survival stratification: confirmed *POLE*mut cases exhibited a 5-year RFS of 97% (82-100), while POLARIX-predicted *POLE*wt outcomes closely matched DNA-confirmed *POLE*wt cases. Although threshold Low yields slightly more false positive results than the Mid and High thresholds (reflected in the lower net benefit), this limitation can be ameliorated because POLARIX requires confirmatory DNA sequencing. Although thresholds Mid and High remain viable alternatives for settings with severe resource constraints, we believe threshold Low is appropriate for both high- and low-resource environments with (centralized) sequencing access.

There are limitations to this study. First, the design assumes that MMRd testing (routinely done for Lynch Syndrome screening) is performed before the use of POLARIX. This might slow down implementation in very low resource settings where immunohistochemistry is not available. Future studies could investigate whether employing POLARIX as a stand-alone *POLE*mut detector in those settings is better than no molecular testing at all. Second, our design choice to limit the use of POLARIX to MMRp EC also implies that *POLE*mut/MMRd double classifiers are not identified as such. Since this is a very small fraction of EC patients (<1%) and the clinical significance of a double *POLE*mut/MMRd classification is not clear, we consider this acceptable^6,23,24^. Third, insufficient data to perform ethnicity-stratified analyses limits the evaluation of potential performance variation across populations.

The next step for POLARIX is validation to confirm clinical performance and probability calibration across diverse patient populations, infrastructures and workflows. Local recalibration within a center may be needed to ensure robustness, reproducibility, and safety across institutional and technical variations^48^. Subsequently, POLARIX should be evaluated in a blinded, prospective study integrated into fully digital pathology workflows using a “silent trial” design^49,50^, where POLARIX predictions are compared with DNA sequencing under pathologist supervision. This approach will generate high-quality evidence of diagnostic safety, efficiency, and concordance, meeting regulatory requirements under frameworks such as the European IVDR and FDA SaMD pathways. After regulatory approval, POLARIX could efficiently allocate *POLE* testing in hospitals with centralized sequencing facilities. H&E slides would be scanned locally, with IHC markers evaluated in parallel. WSIs would be uploaded to a secure cloud platform, where POLARIX computes scores and generates interpretability outputs. Predicted *POLE*wt cases are immediately ruled out, while predicted *POLE*mut cases undergo confirmatory DNA sequencing at regional or national laboratories. This scalable lightweight system reduces unnecessary molecular testing, lowers costs, and maintains clinical safety and can be implemented in both high- and middle-income settings.

By combining diagnostic accuracy, interpretability, and pragmatic clinical design, POLARIX is an example for DL tools aimed not at replacing molecular testing, but at optimizing its allocation. By extending our evaluation beyond discrimination metrics to include calibration, decision utility, and real-world feasibility, we hope to inspire others to explore similar directions that support clinically meaningful AI deployment. As molecular biomarkers increasingly guide therapy, the integration of DL-based screening tools such as POLARIX could enable more equitable access to precision oncology worldwide, changing how molecular information is obtained, interpreted, and applied in routine cancer care.

## Acknowledgements

We are grateful to all patients, investigators, and participating hospitals, as well as the consortia and researchers who curated and shared clinical cohorts contributing to this study. We thank Diantha Terlouw, Leiden University Medical Center, for assistance with interpretation of NGS results, and Natalja ter Haar and Tessa Rutte, Leiden University Medical Center, for excellent technical support. We also thank Damian Linkers for the support on the molecular classification of the UMCU cohort. We acknowledge the AIRMEC Consortium for the exceptional collaboration. The PORTEC-1, PORTEC-2, PORTEC-3, and PORTEC-4a trials were supported by grants from the Dutch Cancer Society (CKTO 90-01, CKTO 2001-04, CKTO 2006-04, UL2011-5336 and UL20, respectively). We acknowledge the PORTEC-3 study group and the TransPORTEC Research Consortium for establishing and maintaining the TransPORTEC biobank. We also acknowledge the Netherlands Cancer Institute for access to their 3D-HISTECH P1000 scanner. We thank Lisa Vermij, Alicia León-Castillo and Ellen Stelloo for the contribution to molecularly classifying the samples. We acknowledge Famke Wakkerman for retrieving additional data for the LUMC cohort and maintaining the collected database. We acknowledge the SHARK team, the computational cluster of the LUMC, for their technical support. This work was supported by The Hanarth Foundation.

## Author Contributions Statement

T.B., N.H., V.K. conceived the study. N.B., L.S., S.A., M.L., J.B.W., E.S., V.K., N.H., T.B. participated in designing the study. T.B., N.H., V.T.H.B.M.S. supervised the study. N.B., S.V.F., R.R., K.G., J.J.J., M.B., C.L.C., R.A.N., S.M.d.B., M.E.P., A.L., G.O., T.J., C.K., D.T., N.K., P.K., L.S., S.K., A.S.V.M.H., M.A.D.H. collected the data. N.B., S.V.F., J.B.W. contributed to code writing, N.B. contributed to model training, N.B. and L.S. contributed to data analysis. J.B.W. prepared the code repository. T.B., N.H., V.K., E.S. contributed to expert review and data interpretation. N.B. designed the figures and drafted the manuscript. N.B., L.S., S.A., M.L., J.B.W., V.K., N.H., T.B., D.C. revised the manuscript. R.R., K.G., J.J.J., M.B., R.A.N., S.M.d.B., A.L., G.O., T.J., C.K., D.T., N.K., M.E.P., P.K., L.S., A.L., S.K., A.S.V.M.H., M.A.D.H., C.L.C. contributed to providing data and tumour material used in this study. All authors had access to the final version of the manuscript and could read and approve. Due to general data protection regulations, verification and access to raw data was limited to N.B., J.B.W., N.H., and T.B. All authors agreed on submitting the manuscript.

## Competing Interests Statement

S.V.F. reports paid consultancy for Tempus AI, Inc. D.C. serves on an advisory board for Merck Sharp & Dohme, received research funding from Cancer Research UK, GSK, Moderna and Veracyte, and speaker fees from GSK, all unrelated to the present study, and declares no personal stock ownership; D.C. is the spouse of an Amgen employee. C.L.C. received grants from the Dutch Cancer Society for the PORTEC-1, -2, -3 and -4a trials and associated translational research unrelated to the present study, and reports institutional research support from Varian and compensation from Merck for data monitoring committee activities, outside the submitted work. L.S. reports participation as an invited speaker for Indica Labs. G.O. received research support from the Fonden af 17-12-1981. A.L. received research funding unrelated to the present study from Owkin, AstraZeneca, Daiichi Sankyo, Zentalis, Genmab, GSK, MSD, Compugen, OSE Immunotherapeutics, Eikon and AbbVie, and consulting fees from Medscape and Clinical Care Options; A.L. also participated in company-sponsored speaker’s bureaus, with fees paid to the institution. S.M.d.B. received research support from Varian Medical Systems and honoraria for lectures from the European School of Oncology, with payments made to the institution, and served as invited speaker and scientific program committee member for ESMO annual meetings (2022–2023), all outside the submitted work. T.B. and A.S.V.M.H. received research funding from the Dutch Cancer Society, with T.B. additionally supported by the Hanarth Fund, unrelated to the present study. V.K. reports sponsored research agreements with Roche and IAG, honoraria or consulting fees from Takeda and Roche, and participation as an invited speaker for Sharing Progress in Cancer Care and Indica Labs, all unrelated to the present study. P.K. reports honoraria from MSD, paid to the institution and used to support the Wellbeing project (2024). E.W.S. reports receiving royalties for a text book ‘Clinical Prediction Models’ from Springer. The other authors declare no competing interests.

## Methods

### Ethics statement

The PORTEC-1, PORTEC-2 (P01.146), PORTEC-3 (P06.031) and PORTEC-4a (P16.054) study protocols were approved by the Medical Ethical Committee Leiden, Den Haag, Delft and the medical ethics committees at participating centers. All study participants of the clinical trials provided informed consent. Studies were conducted in accordance with the principles of the Declaration of Helsinki. Permissions for the retrospective use of the cohorts and a waiver for informed consent was provided for the Transportec High Risk Pilot (B21.065), Medisch Spectrum Twente cohorts (MST, B21.011), Leiden cohort (nWMO-D4-2023-002), Stage-IV cohort (IRBDm20-020; Antoni van Leeuwenhoek), South Africa cohort (HREC: N23/09/119), and UMCU cohort (nWMO-2022-454). Permission for use of the Danish Cohort was provided by the Center for Regional Udvikling, De Videnskabsetiske Komiteer (H-16025909).

### Cohorts

The PORTEC-1 trial^51^ recruited 714 early-stage intermediate risk EC patients from 1990 to 1997 in the Netherlands with the following inclusion criteria: the International Federation of Gynecology and Obstetrics (FIGO) 1988 stage I and grade 1-2 and >50% myometrial invasion; FIGO 1988 stage I and grade 2-3 with <50% invasion. The patients were randomly allocated no adjuvant treatment or external beam radiation therapy. The PORTEC-2 trial^28^ recruited 427 early-stage high-to-intermediate risk EC patients from 2000 to 2006 in the Netherlands with the following inclusion criteria: age over 60 years, FIGO 1988 stage 1B having <50% myometrial invasion, and grade 3; age over 60 years, FIGO 1988 stage 1C having (≥50% myometrial invasion, and grade 1-2; any age, FIGO 1988 stage IIA having endocervical glandular involvement, but not grade 3 with deep invasion. The patients were randomized to receive adjuvant vaginal brachytherapy or external beam radiation therapy. The PORTEC-3 trial^39^ recruited 660 high-risk EC patients from 2006 to 2013 internationally (Australia, Canada, France, the Netherlands, New Zealand, Italy, UK) with the following inclusion criteria: FIGO 2009 stage 1A, endometrioid EC, grade 3, and LVSI; FIGO stage IB, endometrioid EC, grade 3; FIGO 2009 stage II-III and endometrioid EC; FIGO 2009 stage IA-III and serous or clear cell EC. The patients were randomly allocated to receive adjuvant external beam radiation therapy alone or combined radiotherapy and chemotherapy. The PORTEC-4A^29^ trial recruited 569 high-intermediate risk EC patients from 2016 to 2021 internationally with the following inclusion criteria: stage IA (with invasion) and grade 3, stage IB with grade 1 or 2 and age over 60 years, stage IB with grade 1 or 2 with LVSI, stage IB with grade 3 and no LVSI, and stage II with grade 1. The randomization (2:1) was based on the molecular risk profile: the experimental arm consisted of observation for a favorable profile, vaginal brachytherapy for an intermediate risk profile, and external beam radiation therapy for an unfavorable risk profile. The standard arm was adjuvant vaginal brachytherapy. The HR-Pilot^34^ study retrospectively included 116 high-risk EC patients in 2005 from five institutions (*n* = 14 from Institute Gustave Roussy, Villejuif, France; *n* = 14 from the Leiden University Medical Center, The Netherlands, *n* = 46 from the University Medical Center Groningen, The Netherlands, the Netherlands; *n* = 8 from the University College London, United Kingdom and *n* = 34 from St Mary’s Hospital, Manchester, United Kingdom) following the same inclusion criteria as the PORTEC-3 trial. The Medisch Spectrum Twente (MST)-I^30^ cohort consists of 277 prospectively included high-risk EC patients following the PORTEC-3 inclusion criteria and who were also treated with adjuvant radiation therapy between 1987 and 2015 at the MST, Enschede, The Netherlands. The MST-II cohort consists of 170 prospectively included EC patients with FIGO Stage I with an indication for radiotherapy without combination with chemotherapy between 1987 and 2015 at the MST, Enschede, The Netherlands. The Leiden University Medical Center^32^ (LUMC) cohort is a retrospective population cohort of 356 EC patients undergoing surgery at the LUMC in the Netherlands. Patients were included if they were diagnosed with FIGO 2009 stage I-III and were treated with primary surgical treatment at the LUMC. Importantly, patients who were already included in trials or other cohorts used in this study were excluded from this cohort. Patients who underwent neo-adjuvant treatment (either hormonal therapy due to fertility-sparing treatment or chemotherapy due to inoperable disease) were also excluded. The Danish^52^ cohort consists of 450 high-grade EC patients prospectively collected from the Danish gynecological cancer database. The Stage-IV^33^ cohort is a retrospective collection of 165 EC patients with stage IV at diagnosis from five hospitals based in the Netherlands (including Antoni van Leeuwenhoek Hospital, Amsterdam UMC, University Medical Center Utrecht (UMCU), and LUMC). The South-Africa^35^ cohort is a retrospective high-intermediate and high-risk cohort of 148 EC patients diagnosed between 2016 and 2021 treated at Tygerberg Academic Hospital, Cape Town, South Africa, where inclusion followed the following criteria: non-endometrioid EC, grade 3 endometrioid EC, FIGO stage II-III endometrioid EC, grade 1-2 with FIGO stage IB endometrioid EC with no or focal LVSI. The University Medical Center Utrecht (UMCU)-HR^40^ cohort is a retrospective high-risk cohort of 151 patients enrolled between 2011 and 2021 with the following criteria: clinical FIGO stage I-II, grade 3 endometrioid, or serous, clear cell or uterine carcinosarcoma and RALS surgical staging. The Clinical Proteomic Tumor Analysis Consortium Uterine Corpus Endometrial Carcinoma Collection (CPTAC-UCEC)^37,38^ is a public open-source dataset with 122 EC patients, which can be downloaded from The Cancer Imaging Archive (TCIA) at https://www.cancerimagingarchive.net/collection/cptac-ucec/. The Cancer Genome Atlas Uterine Corpus Endometrial Carcinoma (TCGA-UCEC)^36^ is a public dataset with 531 EC patients, which can be downloaded from the cBioPortal at https://www.cbioportal.org/study/summary?id=ucec_tcga_pan_can_atlas_2018.

### Datasets

We collected formalin-fixed, paraffin-embedded (FFPE) tissue tumor material of the hysterectomy endometrial cancer (EC) patients from four randomized trials, eight clinical cohorts, and two public datasets of 4,868 patients (Supplementary Fig. 1). Patients with incomplete or missing molecular class data (*n* = 681) or with MMR-deficient status (*n* = 1,177) were excluded. Hematoxylin and eosin (H&E) whole-slide images (WSIs) were selected by central pathologists and an expert pathologist (TB) based on optimal representation of morphology. For external cohort MST-II, three FFPE blocks all having one representative slide per patient were selected. H&E slides from the randomized trials and clinical cohorts were scanned at 40x magnification on one of three scanners: 3D-Histech P250 (resolution 0.19 µm per pixel), 3D-Histech P480 (resolution 0.19 µm per pixel), and 3D-Histech P1000 (resolution 0.24 µm per pixel) (Supplementary Table 3). A total of 290 patients were excluded if all scans did not meet quality control requirements, which were: (almost) no tumor in the H&E, bad tissue quality, failed scanning, non-hysterectomy slide, no H&E available, or missing data. This exclusion process led to a total of 2,719 patients included in the study (Extended Figure 1, Supplementary Fig. 1). WSIs were preprocessed by segmenting tissue regions. Nonoverlapping square patches of 180 µm edge length were extracted at 40x magnification (0.25 µm/px). All patches were resized to 224 × 224 pixels before feature extraction. Patches with more than 20% white background were excluded. Patches were ImageNet normalized^53^. Patch-level features were extracted using the H-optimus-1 model^25^. The output for each patch was a 1536-dimensional feature vector along with the corresponding patch coordinates.

In total, there were 2,719 patients included in the analyses of this study. From the total, 2,238 were used for the development of the model, while 481 were used as the external test set. The development dataset consisted of three randomized trials, six clinical cohorts, and two public datasets. From the randomized trials, 335 patients of PORTEC-1^27^ (*POLE*mut: *n* = 35 (10.45%)), 277 from PORTEC-2^28^ (*POLE*mut: *n* = 21 (7.58%)), and 386 from PORTEC-4A^29^ (*POLE*mut: *n* = 45 (11.66%)) were included. From the clinical cohorts, we included 87 from the HR-Pilot^34^ (*POLE*mut: *n* = 13 (14.94%)), 168 from Medisch Spectrum Twente (MST)-I^13^ (*POLE*mut: *n* = 12 (7.14%)), 238 from the Leiden University Medical Center (LUMC) cohort (*POLE*mut: *n* = 21 (8.82%)), 272 patients from the Danish^15^ cohort (*POLE*mut: *n* = 30 (11.03%)), 21 from the Stage-IV^16^ cohort (*POLE*mut: *n* = 1 (4.76%)), and 100 patients from the South-Africa^35^ cohort (*POLE*mut: *n* = 6 (6.00%)). From the public datasets, we used 42 from The Clinical Proteomic Tumor Analysis Consortium Uterine Corpus Endometrial Carcinoma Collection (CPTAC-UCEC)^37,38^ (*POLE*mut: *n* = 5 (11.90%)) and 313 from The Cancer Genome Atlas Uterine Corpus Endometrial Carcinoma (TCGA-UCEC)^36^ (*POLE*mut: *n* = 55 (17.57%)). Because of the rarity of *POLE*mut cases, we enriched the development set with an additional 90 *POLE*mut slides (54 from PORTEC-4A, one from MST-I, 29 from LUMC, and six from CPTAC-UCEC) originating from *POLE*mut patients in the model development set. All slides from the same patient were assigned to the same cross-validation fold to prevent leakage. Furthermore, the development set was used to fit the Platt Scaler^54^.

The calibration set consisted of 20% of the development set and was not used in training of the final model. This set was used for early-stopping to mitigate overfitting when training the final model on the remaining 80% of the development set, and for evaluating whether Platt-scaled POLARIX-scores showed better calibration than un-scaled POLARIX-scores.

For the external test set, we included 248 patients from the randomized trial PORTEC-3 (*POLE*mut: *n* = 38 (15.32%)), 111 patients from intermediate-to-high-risk clinical cohort MST-II (*POLE*mut: *n* = 16 (14.41%)), and 122 patients from high-risk cohort UMCU-HR^40^ (*POLE*mut: *n* = 14 (11.48%)). These cohorts were chosen as external test cohorts for their internationality and diversity in risk groups. For all patients, the molecular class was determined using the surrogate marker-based diagnostic algorithm introduced in WHO 2020^55^, yielding *POLE*mut, p53abn, or NSMP as the molecular class.

For external testing, we performed a sample size calculation using the precision method. Details are provided in the Supplement. Briefly, the standard error of the area under the curve (AUC) was approximated using the method described by Hanley and McNeil, using the observed *POLE*mut to *POLE*wt ratio (*r* = 68/481 = 0.165)^56^. Assuming a true AUC of 0.95 and targeting a 95% CI ≥ 0.90, this approach indicated that at least 37 *POLE*mut cases and 221 *POLE*wt cases (total *n* = 257) were required. We therefore pooled the PORTEC-3, MST-II, and UMCU-HR cohorts into a single “Pooled” external test set. All cohort-specific results are reported separately in the Results section and/or in the Supplementary Information.

### POLARIX development and inference

Hyperparameters, network architectures, and foundation model choice (CONCH^57^, H-optimus-1^25^, Virchow2^58^, and UNI-v2^59^) were optimized via five-fold cross-validation, maximizing mean AUC (Supplementary Fig. 2). To address class imbalance in the limited number of *POLE*mut cases, we compared sampling strategies (downsampling, balanced sampling, random sampling) and investigated the use of class-weighted binary cross-entropy loss. The final model was trained on 80% of the dataset using the best-performing hyperparameters and foundation model (H-optimus-1^25^). The Adam optimizer was used with an initial learning rate of 3.0×10^-6^, milestones {5, 15, 25}, gamma of 0.1, weight decay of 1.0×10^-6^. POLARIX was trained on a single NVIDIA Quadro RTX 6000 GPU with 24GB of VRAM for 93 epochs in around 19 hours. The remaining 20% was used as a validation set to inform early stopping and served for calibration via a Platt scaler.

POLARIX combines feature extraction with foundation model H-optimus-1^25^ with a 984K parameter multiple instance learning network with attention pooling that predicts the likelihood of *POLE*mut from H&E WSIs. Input features are 1536-dimensional H-optimus-1^25^ embeddings extracted from 180 µm patches at 40× magnification (0.25 µm/px). Patch embeddings are weighted by a gated attention mechanism and aggregated into a slide-level representation^26^, followed by a fully-connected layer and a sigmoid classifier producing a continuous score between zero and one which is transformed into a probability using a Platt scaler. On average, POLARIX inference on a single WSI takes three minutes for feature extraction with foundation model H-optimus-1^25^ and 0.33 seconds for POLARIX score prediction per slide on a single Quadro RTX 6000 GPU (released in 2018).

### Robustness experiments

In the stain augmentation experiment, POLARIX was tested on the external pooled cohort under simulated changes in stain concentration and color under the Beer-Lambert model of light absorption^60^ (Supplementary Figs. 3-4). We simulated four corner settings, by augmenting WSIs patch-by-patch to reference H&E concentrations and stains extracted from the Pathology Images of Scanners and Mobilephones (PLISM) dataset^61^. Variation in concentration and H&E vector angle in Optical Density (OD) space in three cohorts were reported. This represented the H&E stain similarity, where a lower angle equals a higher similarity, and H&E color variation was based on the first principal component (PC1) value of the cohort’s stain vectors. Intrapatient discordance in the MST-II cohort was estimated by predicting POLARIX-scores for all slides of the patients, as they either two (*n* = 9) or three (*n* = 102) slides originating from different FFPE blocks, and evaluating the absolute maximum difference in POLARIX-scores per patient. For the *POLE* variant analysis, POLARIX-scores were stratified on *POLE* variants and compared for differences.

### Evaluation metrics

Model performance was evaluated for discriminative ability, calibration, classification, and clinical utility^62^. Discrimination was quantified by the AUC. Reliability of predicted probabilities was assessed with calibration analyses, reporting calibration-in-the-large, observed-to-expected event ratio, and calibration plots. Classification performance was evaluated by dichotomizing POLARIX-scores with the thresholds into POLARIX-mutant and -wildtype and reporting sensitivity, specificity, positive predictive value (PPV), negative predictive value (NPV), as well as the test reduction quantified by the true-negative fraction and the missed true *POLE*mut cases quantified by the false-negative fraction. Clinical utility was assessed by a decision-curve analysis (DCA) and reporting the net benefit (NB). All performance evaluations were done on the pooled external test cohort and separate external test cohorts to evaluate POLARIX for fairness.

### Calibration assessment

We assessed model calibration using calibration plots and the calibration metrics O:E ratio, intercept, and slope. The O:E ratio (ideal = 1) measures overall calibration, indicating whether the model predicts too many or too few events on average. The intercept (ideal = 0) evaluates calibration-in-the-large, reflecting systematic over- or underestimation of risk. The slope (ideal = 1) assesses whether predicted probabilities are too extreme, i.e., if high-risk predictions are overestimated and low-risk predictions underestimated. Grouped predictions and observations were compared in lower-risk regimen (0-0.1), intermediate-risk regimen (0.1-0.9) and higher-risk regimen (0.9-1.0).

### Net Benefit computation

Decision curve analysis (DCA) was used to evaluate the clinical utility of POLARIX^41^. It calculates the “Net Benefit” (NB) by using the following formula:

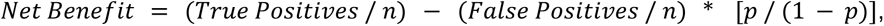

where *n* is the number of patients and *p* is the probability threshold. The formula subtracts the proportion of false positive cases from the proportion of true positive cases, weighted by the relative harm of false positive and false negative cases. This formula estimates the NB per patient, allowing comparison of the model to “NGS test all” or “NGS test none” strategies. A positive NB indicates a net gain in correctly identified cases, while a negative net benefit suggests more harm than benefit. We plotted the decision curve between thresholds 0.00-0.20, which corresponds to the range of probabilities that is suited for a cut-off for a screening system, which finds false positives less harmful than false negatives. NB was calculated at our determined thresholds, Low, Mid, and High, and compared to the “NGS test all” and “NGS test none” strategies. Delta (Δ) Net Benefit is used for the difference in NB, and is positive if the model (POLARIX) is more beneficial than “NGS test all” or “NGS test none”.

### Explainability experiments

Heatmaps of WSIs were generated by first segmenting the tissue and extracting non-overlapping patches. Each patch was represented by its H-optimus-1^25^ embedding and fed into POLARIX, yielding one attention score per patch. Attention scores were standardized (z-transformation) at the slide level and mapped onto a lower-resolution image using the Jet colormap from OpenCV^63^, having red for high attention and blue for low attention. Highly attended patches in slides were extracted using the coordinates of the patches with the highest raw attention scores.

In total, 393 WSIs in cohort PORTEC-3 were annotated by an expert GYN-pathologist (TB) using QuPath^64^ version 0.2.0 for three regions: the tumor region, invasive border, and myometrium (Supplementary Fig. 8). The invasive border was only annotated when the distance between the serosa and the tumor border was more than 1000µm. The regions did not overlap. Approximately 75% of the total number of tumor cells were included in the tumor region. The invasive border was composed of 50/50 tumor and myometrium cells, and was 1000µm wide on both sides. Annotated regions did not include artifacts such as cutting scratches, large areas of necrosis, or tissue folds. This process resulted in 393 cases with tumor region, 351 cases with invasive border annotations, and 355 cases with myometrium annotated.

Proportional attention was computed by softmax transforming the raw attention scores in a WSI to facilitate comparisons between slides. The normalized attention per region was computed by taking the sum of the raw attention scores in the region of interest and dividing by its size in pixels. This way, this score was not dependent on the size of the region, limiting human-introduced bias, and enabling comparisons of attention between regions.

To count the number of inflammatory cells and to compute the nuclear atypia score, we used CellViT^++65^ with the pre-trained PanNuke^66^ weights to classify detected cells in WSIs (Supplementary Fig. 9). The PanNuke^66^ classification includes connective, epithelial, inflammatory, non-nuclei, and neoplastic cells. All cells on a WSI were segmented, classified, and morphometrics were calculated, including nuclei area size, sphericity, and convexity based on their contours. These results were aggregated at the slide-level per cell type using the median and IQR.

The mitotic density score was determined by searching the location with the highest density of automatically detected mitotic figures within a disk of area 1mm^2^ in a given WSI. These mitotic figures were detected using a deep learning model^67^ that was specifically trained and validated for the detection of mitotic figures in EC^68^. Tertiary lymphoid structures were counted on WSI level using immunohistochemistry staining with L1CAM^44^. CD8 Density was determined with the average log^2^ transformed density of CD8^+^ lymphocytes in the tumor and stroma in a TMA^44^. The fraction of inflammatory cells was computed using the aggregated results of the classifications of the CellViT^++65^ analysis by dividing the inflammatory cell count by the total number of cells for each WSI. The median area of tumor cells was computed using the aggregated area metric of the CellViT^++^ analysis. We define the nuclear atypia score as the average of non-convexity and non-sphericity (Supplement Information 4.3). The metrics are aggregated results using the median from the CellViT^++^ analysis on WSI level.

### Statistical analysis

AUC metrics and classification metrics were evaluated using *n* = 1000 times bootstrapping to report mean with 95% confidence interval. Platt scaling was used to transform scores to probabilities. POLARIX-score calibration was assessed with flexible calibration curves using “rcs” as the smoother, confidence levels (CL), and three pre-specified probability bins (0-0.1, 0.1-0.9, 0.9-1). In each bin, we plotted the mean predicted probability against the observed event rate (triangular points) with 95% confidence (Wilson) intervals. A multiple linear regression was performed to analyze associations between clinicopathological variables and the POLARIX-score. Fisher 2×2 exact tests were performed to compare clinicopathological characteristic distributions between prediction groups. Mann-Whitney-Wilcoxon test two-sided tests were performed to compare POLARIX-score distributions between *POLE* variants and morphologic feature distributions between prediction groups. Spearman correlation analysis was performed for the correlation between two continuous variables. Statistical significance was accepted with *p*-values below 0.050. Multiple hypothesis testing was corrected for using the Bonferroni method. Kaplan-Meier analyses were performed to analyze clinical outcomes with patient subgroup stratification, where the log-rank test examined the difference between outcomes.

### Software

POLARIX was developed in Python version 3.12.9 using PyTorch^69^ version 2.7.0. All experiments were performed using Python except for the linear regression and calibration assessments which were done using R. The package OpenSlide^70^ was used for pre-processing the WSIs. Matplotlib and Seaborn^71^ were used to create plots. Adobe Illustrator was used to edit figures. The calibration plots were made in R Studio using the CalibrationCurves package and the “val.prob.ci.2” function. Decision curve analysis (DCA)^41^ was performed using the package “dcurves” in Python. Kaplan-Meier analysis was done using Python package Lifelines.

## Data Availability

All tumor material and datasets that were used in this study, except for publicly available cohort CPTAC-UCEC and TCGA-UCEC, are not publicly available due to restrictions by privacy laws. The PORTEC study group and international TransPORTEC consortium hold the material and data from PORTEC-1, PORTEC-2, PORTEC-3, PORTEC-4A, MST-I, MST-II, and HR-Pilot. Data and materials from the LUMC and Stage-IV cohorts are held by N.H and T.B., the Danish cohort are held by G.O., the South Africa cohort are held by R.R., the UMCU-HR cohort are held by K.G. Sharing the data and material can only be done on request within 15 years of the date of publication of this Article, and includes a scientific proposal addressed to the corresponding author. Subject to ethical consent and depending on the specific research proposal, the TransPORTEC consortium, PORTEC study group, G.O., R.R., or K.G. will determine how long, when, and how the requested data can be made available. CPTAC-UCEC data can be downloaded from https://www.cancerimagingarchive.net/collection/cptac-ucec/. TCGA-UCEC data can be downloaded from https://www.cbioportal.org/study/summary?id=ucec_tcga_pan_can_atlas_2018.

## Code Availability

Code used in this study is available via GitHub at https://github.com/AIRMEC/POLARIX.

## Notes

### Clinical Trial

PORTEC-2 (NCT00376844), PORTEC-3 (NCT00411138), PORTEC-4a (NCT03469674)

### Author Declarations

The Medical Ethical Committee Leiden, Den Haag, Delft and the medical ethics committees at participating centers gave ethical approval for the PORTEC-1 (No protocol number available, 90's trial), PORTEC-2 (P01.146), PORTEC-3 (P06.031), and PORTEC-4a (P16.054) study protocols for this work. All study participants of the clinical trials provided informed consent. Studies were conducted in accordance with the principles of the Declaration of Helsinki. Ethics committee Leiden, Den Haag, Delft and the medical ethics committees at participating centers gave ethical approval for the retrospective use of the cohorts and a waiver for informed consent was provided for the Transportec High Risk Pilot (B21.065), Medisch Spectrum Twente cohorts (MST, B21.011), Leiden cohort (nWMO‐D4‐2023‐002), Stage-IV cohort (IRBDm20-020; Antoni van Leeuwenhoek), South Africa cohort (HREC: N23/09/119), and UMCU cohort (nWMO-2022-454) for this work. The Center for Regional Udvikling, De Videnskabsetiske Komiteer, gave ethical approval for use of the Danish Cohort for this work (H-16025909).

