## Supplementary Information for "Deep Learning-Based Screening for *POLE* mutations on Histopathology Slides in Endometrial Cancer"

#### Table of Contents

**1. Data** **2**
1.1. Cohort explanation 2
1.2. Patient and case exclusion 3
1.3. Slide selection and scanning 9
**2. Developing POLARIX** **9**
2.1. Foundation model selection and hyperparameter optimization 9
**3. Performance experiments** **10**
3.1. Robustness checks 10
3.2. Fitting the Platt scaler for calibration 11
3.3. Calibration assessment 12
3.4. Classification accuracy 13
3.5. Net Benefit computation 15
**4. Explainability experiments** **16**
4.1. Clinicopathological correlates of POLARIX-scores 16
4.2. Attention analyses 16
4.3. CellViT++ deployment and feature analysis 17
4.4. Misclassification Review in PORTEC-3 18
**4.5. Clinical outcome experiments** **22**
**5. Sample size calculation** **22**

### 1. Data

#### 1.1. Cohort explanation

##### *Randomized trials*

The PORTEC-1 trial[1] recruited 714 patients with early-stage intermediate risk EC from 1990 to 1997 in the Netherlands with the following inclusion criteria: the International Federation of Gynecology and Obstetrics (FIGO) 1988 stage I and grade 1-2 and >50% myometrial invasion; FIGO 1988 stage I and grade 2-3 with <50% invasion. The patients were randomly allocated to no adjuvant treatment or external beam radiation therapy.

The PORTEC-2 trial[2] recruited 427 patients with early-stage high-intermediate risk EC from 2000 to 2006 in the Netherlands with the following inclusion criteria: age over 60 years, FIGO 1988 stage 1B having <50% myometrial invasion, and grade 3; age over 60 years, FIGO 1988 stage 1C having (≥50% myometrial invasion, and grade 1-2; any age, FIGO 1988 stage IIA having endocervical glandular involvement, but not grade 3 with deep invasion. The patients were randomized to receive adjuvant vaginal brachytherapy or external beam radiation therapy.

The PORTEC-3 trial[3] recruited 660 patients with high-risk EC from 2006 to 2013 internationally (Australia and New Zealand, Canada, France, the Netherlands, Italy, the UK) with the following inclusion criteria: FIGO 2009 stage 1A, endometrioid EC, with grade 3 and/or lymphovascular space invasion (LVSI); FIGO stage IB, endometrioid EC grade 3; FIGO 2009 stage II-III and endometrioid EC; FIGO 2009 stage IA-III with serous or clear cell type EC. The patients were randomly allocated to receive adjuvant external beam radiation therapy alone or combined radiotherapy with concurrent and adjuvant chemotherapy.

The PORTEC-4A trial[4] recruited 564 patients with high-intermediate risk EC from 2016 to 2021 internationally with the following inclusion criteria: stage IA (with invasion) and grade 3 stage IB with grade 1 or 2 and age over 60 years and/or with LVSI stage IB with grade 3 and no LVSI and stage II with grade 1. The randomization (2:1) allocated patients to treatment based on the molecular risk profile or standard adjuvant vaginal brachytherapy. Adjuvant treatment in the molecular profile arm consisted of observation for a favorable profile, vaginal brachytherapy for intermediate profile, and external beam radiation therapy for an unfavorable profile.

##### *Clinical cohorts*

The HR-Pilot[5] study retrospectively included 116 high-risk EC patients in 2005 from five institutions (N=14 from Institute Gustave Roussy, Villejuif, France; N=14 from the Leiden University Medical Center, The Netherlands, N=46 from the University Medical Center Groningen, The Netherlands, the Netherlands; N=8 from the University College London, United Kingdom and N=34 from St Mary's Hospital, Manchester, United Kingdom) following the same inclusion criteria as the PORTEC-3 trial.

The Medisch Spectrum Twente (MST)-HR[6] cohort consists of 277 prospectively included high-risk EC patients following the PORTEC-3 inclusion criteria and were treated with adjuvant radiation therapy between 1987 and 2015 at the MST, Enschede, The Netherlands.

The MST-II cohort consists of 170 prospectively included EC patients with FIGO Stage I with an indication for radiotherapy without combination with chemotherapy between 1987 and 2015 at the MST, Enschede, The Netherlands.

The Leiden University Medical Center (LUMC)[7] cohort is a retrospective population cohort of 349 EC patients undergoing surgery at the LUMC in the Netherlands, with an additional enrichment of seven POLEmut EC consultation cases making a total of 356 patients. Patients were included if they were diagnosed with FIGO 2009 stage I-III and were treated with primary surgical treatment at the LUMC.

Importantly, patients who were already included in trials or other cohorts used in this study were excluded from this cohort. Patients who underwent neo-adjuvant treatment (either hormonal therapy due to fertility-sparing treatment or chemotherapy due to inoperable disease) were also excluded.

The Danish[8] cohort consists of 450 high-grade EC patients prospectively collected from the Danish gynecological cancer database between the years 2005 and 2012.

The Stage-IV[9] cohort is a retrospective collection of 165 EC patients with stage IV at diagnosis from five hospitals based in the Netherlands.

The South-Africa[10] cohort is a retrospective high-intermediate and high-risk cohort of 148 EC patients diagnosed between 2016 and 2021 treated at Tygerberg Academic Hospital, Cape Town, South Africa, where inclusion followed the following criteria: 1) non-endometrioid EC, 2) grade 3 endometrioid EC, 3) FIGO stage II-III endometrioid EC, 4) grade 1-2 with FIGO stage IB endometrioid EC with no or focal LVSI.

The University Medical Center Utrecht (UMCU)-HR[11] cohort is a retrospective high-risk cohort of 151 patients enrolled between 2011 and 2021 with the following criteria: 1) clinical FIGO stage I-II, 2) grade 3 endometrioid, or serous, clear cell or uterine carcinosarcoma and 3) RALS surgical staging. The pathology report was used for pathology variables, and not revised in research setting.

*Public datasets*

The Clinical Proteomic Tumor Analysis Consortium Uterine Corpus Endometrial Carcinoma Collection (CPTAC-UCEC)[12,13] is a public open-source dataset with 122 EC patients, which can be downloaded from The Cancer Imaging Archive (TCIA) at https://www.cancerimagingarchive.net/collection/cptac-ucec/.

The Cancer Genome Atlas Uterine Corpus Endometrial Carcinoma (TCGA-UCEC)[14] is a public dataset with 531 EC patients, which can be downloaded from the cBioPortal at https://www.cbioportal.org/study/summary?id=ucec_tcga_pan_can_atlas_2018.

#### 1.2. Patient and case exclusion

*Representative hysterectomy WSIs*

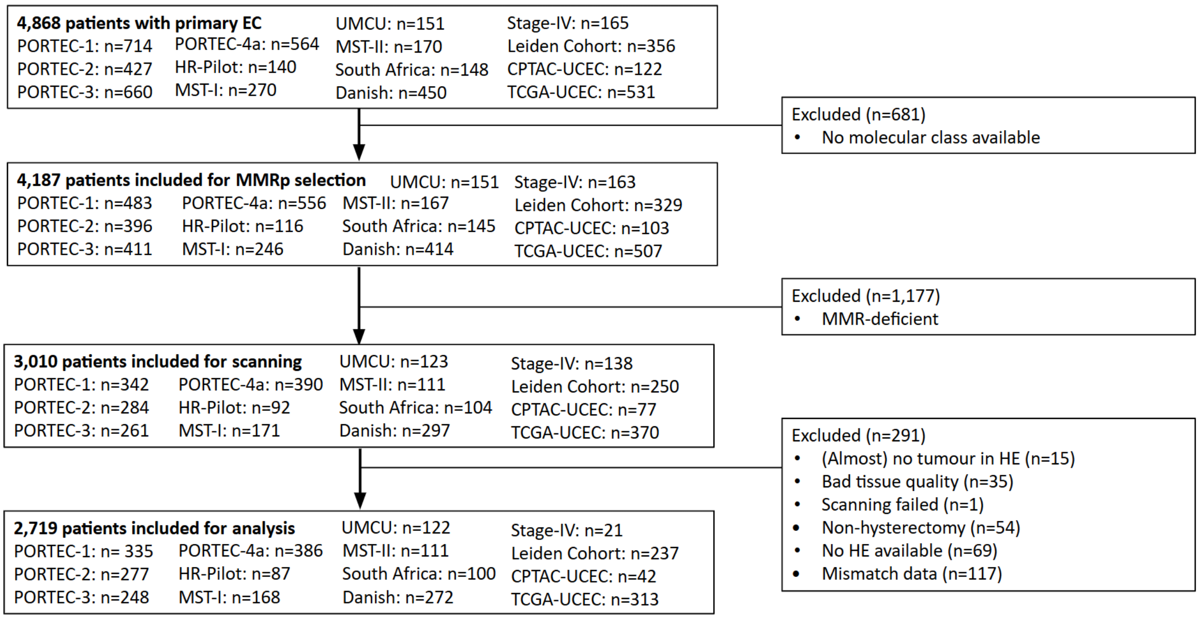

**Figure 1.** CONSORT of the data inclusion process. At starting point, data from 4,868 patients with primary EC were collected from PORTEC-1, PORTEC-2, PORTEC-3, PORTEC-4A, MST-I, MST-II, HR-Pilot, Danish, LUMC, South Africa, UMCU-HR, Stage-IV, CPTAC-UCEC, and TCGA-UCEC. First, 681 patients were excluded due to missing or incomplete molecular class data. Next, we excluded all MMR-deficient patients, which was a total of 1,177 patients. The remaining 3,010 patients were included for scanning. Our expert pathologist reviewed all scans and excluded 291 patients, specifically on 1) (almost) no tumor in the H&E (n=15), 2) bad tissue quality (defined by very extensive necrosis, severe cutting artifacts or tumor tissue with extensive discohesive tumor cells due to poor fixation; n=35), 3) failed scanning (n=1), 4) non-hysterectomy H&E (n=54), 5) no H&E available (n=69), 6) missing data (n=117). These exclusion criteria led to a total of 2,719 patients included for analysis with at least one representative H&E WSI of the hysterectomy of the primary EC. Patient characteristics of the included data are depicted in Table 1-2.

**Table 1.** Patient characteristics of the datasets for development of the prediction model for patients with endometrial cancer.

| | **PORTEC-1 (N=335)** | **PORTEC-2 (N=277)** | **PORTEC-4a (N=386)** | **MST-I cohort (N=168)** | **HRpilot (N=87)** | **Danish (N=272)** | **Leiden cohort (N=237)** | **South-Africa cohort (N=100)** | **Stage IV (N=21)** | **CPTAC (N=42)** | **TCGA (N=313)** | **Overall (N=2238)** |
|---|---|---|---|---|---|---|---|---|---|---|---|---|
| **Age (years)** | | | | | | | | | | | | |
| Mean (SD) | 66.2 (8.54) | 69.7 (7.05) | 68.7 (7.68) | 68.6 (10.4) | 65.4 (11.3) | 70.8 (9.94) | 65.2 (10.6) | 65.8 (9.75) | 66.2 (9.50) | NA (NA) | 63.9 (11.4) | 67.4 (9.69) |
| Median [Min, Max] | 67.0 [41.0, 90.0] | 69.0 [52.0, 88.0] | 69.0 [36.0, 90.0] | 70.0 [25.0, 92.0] | 67.2 [21.0, 85.0] | 70.0 [38.6, 94.8] | 66.0 [29.0, 90.0] | 66.0 [27.0, 92.0] | 68.4 [33.0, 79.0] | NA [NA, NA] | 64.0 [31.0, 90.0] | 68.0 [21.0, 94.8] |
| Missing | 0 (0%) | 0 (0%) | 3 (0.8%) | 0 (0%) | 0 (0%) | 2 (0.7%) | 30 (12.7%) | 2 (2.0%) | 0 (0%) | 42 (100%) | 3 (1.0%) | 82 (3.7%) |
| **Stage** | | | | | | | | | | | | |
| I | 0 (0%) | 0 (0%) | 0 (0%) | 0 (0%) | 0 (0%) | 0 (0%) | 0 (0%) | 0 (0%) | 0 (0%) | 0 (0%) | 15 (4.8%) | 15 (0.7%) |
| IA | 139 (41.5%) | 40 (14.4%) | 42 (10.9%) | 17 (10.1%) | 8 (9.2%) | 113 (41.5%) | 119 (50.2%) | 36 (36.0%) | 0 (0%) | 0 (0%) | 90 (28.8%) | 604 (27.0%) |
| IB | 196 (58.5%) | 233 (84.1%) | 327 (84.7%) | 44 (26.2%) | 22 (25.3%) | 59 (21.7%) | 40 (16.9%) | 11 (11.0%) | 0 (0%) | 0 (0%) | 80 (25.6%) | 1012 (45.2%) |
| II | 0 (0%) | 2 (0.7%) | 17 (4.4%) | 45 (26.8%) | 18 (20.7%) | 17 (6.3%) | 15 (6.3%) | 19 (19.0%) | 0 (0%) | 0 (0%) | 33 (10.5%) | 166 (7.4%) |
| III | 0 (0%) | 0 (0%) | 0 (0%) | 0 (0%) | 0 (0%) | 0 (0%) | 0 (0%) | 0 (0%) | 0 (0%) | 0 (0%) | 1 (0.3%) | 1 (0.0%) |
| IIIA | 0 (0%) | 1 (0.4%) | 0 (0%) | 38 (22.6%) | 16 (18.4%) | 5 (1.8%) | 17 (7.2%) | 6 (6.0%) | 0 (0%) | 0 (0%) | 25 (8.0%) | 108 (4.8%) |

| | | | | | | | | | | | | |
|---|---|---|---|---|---|---|---|---|---|---|---|---|
| IIIB | 0 (0%) | 1 (0.4%) | 0 (0%) | 14 (8.3%) | 1 (1.1%) | 8 (2.9%) | 4 (1.7%) | 0 (0%) | 0 (0%) | 0 (0%) | 2 (0.6%) | 30 (1.3%) |
| IIIC | 0 (0%) | 0 (0%) | 0 (0%) | 10 (6.0%) | 12 (13.8%) | 39 (14.3%) | 12 (5.1%) | 23 (23.0%) | 0 (0%) | 0 (0%) | 48 (15.3%) | 144 (6.4%) |
| IV | 0 (0%) | 0 (0%) | 0 (0%) | 0 (0%) | 9 (10.3%) | 29 (10.7%) | 0 (0%) | 0 (0%) | 21 (100%) | 0 (0%) | 19 (6.1%) | 78 (3.5%) |
| Missing | 0 (0%) | 0 (0%) | 0 (0%) | 0 (0%) | 1 (1.1%) | 2 (0.7%) | 30 (12.7%) | 5 (5.0%) | 0 (0%) | 42 (100%) | 0 (0%) | 80 (3.6%) |
| **Histotype** | | | | | | | | | | | | |
| EEC | 331 (98.8%) | 268 (96.8%) | 380 (98.4%) | 111 (66.1%) | 63 (72.4%) | 96 (35.3%) | 156 (65.8%) | 33 (33.0%) | 7 (33.3%) | 0 (0%) | 220 (70.3%) | 1665 (74.4%) |
| SEC | 3 (0.9%) | 9 (3.2%) | 2 (0.5%) | 20 (11.9%) | 11 (12.6%) | 113 (41.5%) | 36 (15.2%) | 35 (35.0%) | 8 (38.1%) | 6 (14.3%) | 77 (24.6%) | 320 (14.3%) |
| CCC | 1 (0.3%) | 0 (0%) | 0 (0%) | 10 (6.0%) | 13 (14.9%) | 31 (11.4%) | 14 (5.9%) | 5 (5.0%) | 5 (23.8%) | 1 (2.4%) | 0 (0%) | 1665 (74.4%) |
| Carcinosarcoma | 0 (0%) | 0 (0%) | 2 (0.5%) | 23 (13.7%) | 0 (0%) | 14 (5.1%) | 12 (5.1%) | 27 (27.0%) | 0 (0%) | 0 (0%) | 0 (0%) | 78 (3.5%) |
| Un-dedifferentiated | 0 (0%) | 0 (0%) | 0 (0%) | 4 (2.4%) | 0 (0%) | 9 (3.3%) | 2 (0.8%) | 0 (0%) | 1 (4.8%) | 0 (0%) | 1 (0.3%) | 17 (0.8%) |
| Other | 0 (0%) | 0 (0%) | 2 (0.5%) | 0 (0%) | 0 (0%) | 9 (3.3%) | 6 (2.5%) | 0 (0%) | 0 (0%) | 0 (0%) | 15 (4.8%) | 32 (1.4%) |
| Missing | 0 (0%) | 0 (0%) | 0 (0%) | 0 (0%) | 0 (0%) | 0 (0%) | 11 (4.6%) | 0 (0%) | 0 (0%) | 35 (83.3%) | 0 (0%) | 46 (2.1%) |
| **Grade** | | | | | | | | | | | | |
| Grade 1-2 | 279 (83.3%) | 249 (89.9%) | 335 (86.8%) | 72 (42.9%) | 12 (13.8%) | 21 (7.7%) | 141 (59.5%) | 19 (19.0%) | 2 (9.5%) | 0 (0%) | 132 (42.2%) | 1262 (56.4%) |
| Grade 3 | 56 (16.7%) | 28 (10.1%) | 51 (13.2%) | 96 (57.1%) | 75 (86.2%) | 251 (92.3%) | 85 (35.9%) | 81 (81.0%) | 19 (90.5%) | 7 (16.7%) | 181 (57.8%) | 930 (41.6%) |
| Missing | 0 (0%) | 0 (0%) | 0 (0%) | 0 (0%) | 0 (0%) | 0 (0%) | 11 (4.6%) | 0 (0%) | 0 (0%) | 35 (83.3%) | 0 (0%) | 46 (2.1%) |
| **LVSI** | | | | | | | | | | | | |

| | | | | | | | | | | | | |
|---|---|---|---|---|---|---|---|---|---|---|---|---|
| Focal or absent | 323 (96.4%) | 266 (96.0%) | 365 (94.6%) | 134 (79.8%) | 34 (39.1%) | 235 (86.4%) | 165 (69.6%) | 67 (67.0%) | 0 (0%) | 0 (0%) | 0 (0%) | 1589 (71.0%) |
| Present | 12 (3.6%) | 11 (4.0%) | 21 (5.4%) | 34 (20.2%) | 36 (41.4%) | 37 (13.6%) | 39 (16.5%) | 27 (27.0%) | 0 (0%) | 0 (0%) | 0 (0%) | 217 (9.7%) |
| Missing | 0 (0%) | 0 (0%) | 0 (0%) | 0 (0%) | 17 (19.5%) | 0 (0%) | 33 (13.9%) | 6 (6.0%) | 21 (100%) | 42 (100%) | 313 (100%) | 432 (19.3%) |
| **(Neo) Adjuvant treatment** | | | | | | | | | | | | |
| None | 177 (52.8%) | 1 (0.4%) | 162 (42.0%) | 0 (0%) | 8 (9.2%) | 179 (65.8%) | 81 (34.2%) | 14 (14.0%) | 0 (0%) | 0 (0%) | 0 (0%) | 622 (27.8%) |
| VBT | 0 (0%) | 142 (51.3%) | 201 (52.1%) | 16 (9.5%) | 0 (0%) | 0 (0%) | 35 (14.8%) | 27 (27.0%) | 0 (0%) | 0 (0%) | 0 (0%) | 421 (18.8%) |
| EBRT | 158 (47.2%) | 134 (48.4%) | 23 (6.0%) | 137 (81.5%) | 40 (46.0%) | 21 (7.7%) | 25 (10.5%) | 23 (23.0%) | 0 (0%) | 0 (0%) | 0 (0%) | 561 (25.1%) |
| CT | 0 (0%) | 0 (0%) | 0 (0%) | 0 (0%) | 1 (1.1%) | 64 (23.5%) | 7 (3.0%) | 3 (3.0%) | 0 (0%) | 0 (0%) | 0 (0%) | 75 (3.4%) |
| CT+EBRT | 0 (0%) | 0 (0%) | 0 (0%) | 11 (6.5%) | 2 (2.3%) | 6 (2.2%) | 10 (4.2%) | 5 (5.0%) | 0 (0%) | 0 (0%) | 0 (0%) | 34 (1.5%) |
| VBT+EBRT+CT | 0 (0%) | 0 (0%) | 0 (0%) | 0 (0%) | 7 (8.0%) | 0 (0%) | 8 (3.4%) | 2 (2.0%) | 0 (0%) | 0 (0%) | 0 (0%) | 17 (0.8%) |
| VBT+EBRT | 0 (0%) | 0 (0%) | 0 (0%) | 0 (0%) | 10 (11.5%) | 0 (0%) | 12 (5.1%) | 9 (9.0%) | 0 (0%) | 0 (0%) | 0 (0%) | 31 (1.4%) |
| VBT+CT | 0 (0%) | 0 (0%) | 0 (0%) | 2 (1.2%) | 0 (0%) | 0 (0%) | 0 (0%) | 1 (1.0%) | 0 (0%) | 0 (0%) | 0 (0%) | 3 (0.1%) |
| Debulking alone | 0 (0%) | 0 (0%) | 0 (0%) | 0 (0%) | 0 (0%) | 0 (0%) | 0 (0%) | 0 (0%) | 2 (9.5%) | 0 (0%) | 0 (0%) | 2 (0.1%) |
| Neoadjuvant CT + Debulking | 0 (0%) | 0 (0%) | 0 (0%) | 0 (0%) | 0 (0%) | 0 (0%) | 0 (0%) | 0 (0%) | 8 (38.1%) | 0 (0%) | 0 (0%) | 8 (0.4%) |
| Neoadjuvant CT + Debulking + CT | 0 (0%) | 0 (0%) | 0 (0%) | 0 (0%) | 0 (0%) | 0 (0%) | 0 (0%) | 0 (0%) | 4 (19.0%) | 0 (0%) | 0 (0%) | 4 (0.2%) |

| | | | | | | | | | | | | |
|---|---|---|---|---|---|---|---|---|---|---|---|---|
| Missing | 0 (0%) | 0 (0%) | 0 (0%) | 2 (1.2%) | 19 (21.8%) | 2 (0.7%) | 59 (24.9%) | 16 (16.0%) | 7 (33.3%) | 42 (100%) | 313 (100%) | 460 (20.6%) |
| **Molecular Markers** | | | | | | | | | | | | |
| POLE wt + p53wt | 260 (77.6%) | 227 (81.9%) | 322 (83.4%) | 88 (52.4%) | 42 (48.3%) | 57 (21.0%) | 146 (61.6%) | 24 (24.0%) | 7 (33.3%) | 26 (61.9%) | 132 (42.2%) | 1331 (59.5%) |
| POLE mut + p53wt | 29 (8.7%) | 20 (7.2%) | 35 (9.1%) | 8 (4.8%) | 11 (12.6%) | 22 (8.1%) | 21 (8.9%) | 4 (4.0%) | 1 (4.8%) | 4 (9.5%) | 52 (16.6%) | 207 (9.2%) |
| POLE wt + p53abn | 40 (11.9%) | 29 (10.5%) | 19 (4.9%) | 68 (40.5%) | 32 (36.8%) | 185 (68.0%) | 69 (29.1%) | 70 (70.0%) | 13 (61.9%) | 11 (26.2%) | 126 (40.3%) | 662 (29.6%) |
| POLE mut + p53abn | 6 (1.8%) | 1 (0.4%) | 10 (2.6%) | 4 (2.4%) | 2 (2.3%) | 8 (2.9%) | 1 (0.4%) | 2 (2.0%) | 0 (0%) | 1 (2.4%) | 3 (1.0%) | 38 (1.7%) |

**Table 2.** Patient characteristics of external test cohorts including 481 patients with endometrial cancer.

| | **PORTEC-3 (N=248)** | **MST-II cohort (N=111)** | **UMCU (N=122)** | **Overall (N=481)** |
|---|---|---|---|---|
| **Age (years)** | | | | |
| Mean (SD) | 61.4 (9.15) | 68.5 (7.36) | 67.8 (7.77) | 64.7 (9.07) |
| Median [Min, Max] | 62.4 [26.7, 80.5] | 68.0 [50.0, 84.0] | 69.0 [42.0, 83.0] | 65.5 [26.7, 84.0] |
| **Stage** | | | | |
| I | 0 (0%) | 0 (0%) | 0 (0%) | 0 (0%) |
| IA | 34 (13.7%) | 23 (20.7%) | 103 (84.4%) | 160 (33.3%) |
| IB | 41 (16.5%) | 88 (79.3%) | 8 (6.6%) | 137 (28.5%) |
| II | 68 (27.4%) | 0 (0%) | 11 (9.0%) | 79 (16.4%) |
| III | 0 (0%) | 0 (0%) | 0 (0%) | 0 (0%) |
| IIIA | 34 (13.7%) | 0 (0%) | 0 (0%) | 34 (7.1%) |
| IIIB | 14 (5.6%) | 0 (0%) | 0 (0%) | 14 (2.9%) |
| IIIC | 57 (23.0%) | 0 (0%) | 0 (0%) | 57 (11.9%) |
| IV | 0 (0%) | 0 (0%) | 0 (0%) | 0 (0%) |
| Missing | 0 (0%) | 0 (0%) | 0 (0%) | 0 (0%) |
| **Histotype** | | | | |
| EEC | 176 (71.0%) | 98 (88.3%) | 37 (30.3%) | 311 (64.7%) |

| | | | | |
|---|---|---|---|---|
| SEC | 53 (21.4%) | 7 (6.3%) | 43 (35.2%) | 103 (21.4%) |
| CCC | 12 (4.8%) | 0 (0%) | 11 (9.0%) | 23 (4.8%) |
| Carcinosarcoma | 0 (0%) | 3 (2.7%) | 26 (21.3%) | 29 (6.0%) |
| Un-dedifferentiated | 3 (1.2%) | 1 (0.9%) | 4 (3.3%) | 8 (1.7%) |
| Other | 3 (1.2%) | 1 (0.9%) | 1 (0.8%) | 5 (1.0%) |
| Missing | 1 (0.4%) | 1 (0.9%) | 0 (0%) | 2 (0.4%) |
| **Grade** | | | | |
| Grade 1-2 | 102 (41.1%) | 90 (81.1%) | 5 (4.1%) | 197 (41.0%) |
| Grade 3 | 145 (58.5%) | 21 (18.9%) | 117 (95.9%) | 283 (58.8%) |
| Missing | 1 (0.4%) | 0 (0%) | 0 (0%) | 1 (0.2%) |
| **LVSI** | | | | |
| Present | 148 (59.7%) | 3 (2.7%) | 4 (3.3%) | 155 (32.2%) |
| Focal or absent | 100 (40.3%) | 106 (95.5%) | 117 (95.9%) | 323 (67.2%) |
| Missing | 0 (0%) | 2 (1.8%) | 1 (0.8%) | 3 (0.6%) |
| **(Neo-) Adjuvant Treatment** | | | | |
| None | 0 (0%) | 0 (0%) | 0 (0%) | 0 (0%) |
| VBT | 0 (0%) | 44 (39.6%) | 0 (0%) | 44 (9.1%) |
| EBRT | 116 (46.8%) | 66 (59.5%) | 0 (0%) | 182 (37.8%) |
| CT | 0 (0%) | 0 (0%) | 0 (0%) | 0 (0%) |
| CT+EBRT | 132 (53.2%) | 0 (0%) | 0 (0%) | 132 (27.4%) |
| VBT+EBRT+CT | 0 (0%) | 0 (0%) | 0 (0%) | 0 (0%) |
| VBT+EBRT | 0 (0%) | 0 (0%) | 0 (0%) | 0 (0%) |
| VBT+CT | 0 (0%) | 0 (0%) | 0 (0%) | 0 (0%) |
| Missing | 0 (0%) | 1 (0.9%) | 122 (100%) | 123 (25.6%) |
| **Molecular Markers** | | | | |
| POLEwt + p53wt | 120 (48.4%) | 82 (73.9%) | 21 (17.2%) | 223 (46.4%) |
| POLEmut + p53wt | 32 (12.9%) | 13 (11.7%) | 6 (4.9%) | 51 (10.6%) |
| POLEwt + p53abn | 90 (36.3%) | 13 (11.7%) | 87 (71.3%) | 190 (39.5%) |
| POLEmut + p53abn | 6 (2.4%) | 3 (2.7%) | 8 (6.6%) | 17 (3.5%) |

#### 1.3. Slide selection and scanning

**Table 3.** Scanner information per cohort. *Exact number not known.

| Cohort | Scanner | Objective magnification | mpxx/mpxy [µm per pixel] | Output resolution |
|---|---|---|---|---|

| | | | | |
|---|---|---|---|---|
| PORTEC-1 | 3D-Histech P250 and P1000 | 40x | 0.194475/0.194475 and 0.249081/0.249081 | 51.4205 and 40.1725 |
| PORTEC-2 | 3D-Histech P250 and P1000 | 40x | 0.194475/0.194475 and 0.249081/0.249081 | 51.4205 and 40.1725 |
| PORTEC-3 | 3D-Histech P250 and P1000 | 40x | 0.194475/0.194475 and 0.249081/0.249081 | 51.4205 and 40.1725 |
| PORTEC-4A | 3D-Histech P480 | 40x | 0.194475/0.194475 | 51.4205 |
| MST-I | 3D-Histech P250 and P1000 | 40x | 0.194475/0.194475 and 0.249081/0.249081 | 51.4205 and 40.1725 |
| MST-II | 3D-Histech P480 | 40x | 0.194475/0.194475 | 51.4205 |
| LUMC | 3D-Histech P1000 | 40x | 0.249081/0.249081 | 40.1725 |
| Danish | 3D-Histech P250 and P1000 | 40x | 0.194475/0.194475 and 0.249081/0.249081 | 51.4205 and 40.1725 |
| HR-Pilot | 3D-Histech P250 and P1000 | 40x | 0.194475/0.194475 and 0.249081/0.249081 | 51.4205 and 40.1725 |
| Stage-IV | 3D-Histech P480 | 40x | 0.194475/0.194475 | 51.4205 |
| South-Africa | 3D-Histech P480 | 40x | 0.194475/0.194475 | 51.4205 |
| UMCU-HR | 3D-Histech P480 | 40x | 0.194475/0.194475 | 51.4205 |
| CPTAC-UCEC | - | 20x | ~0.494/~0.494 | - |
| TCGA-UCEC | - | 40x | ~0.2505/~0.2505* | ~39.92* |

#### 2. Developing POLARIX

##### 2.1. Foundation model selection and hyperparameter optimization

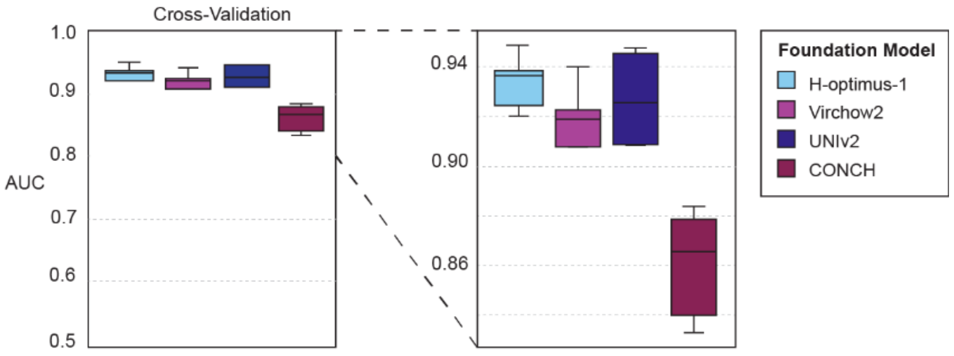


**Figure 2.** Cross-validation results comparing foundation models. We performed five-fold cross-validation to evaluate foundation models for feature extraction from whole-slide images (WSIs). The same POLARIX network architecture was used across all experiments, while varying the foundation model (CONCH[15], H-optimus-1[16], Virchow2[17], and UNI-v2[18]). Among the foundation models evaluated, H-optimus-1[16] achieved the highest mean AUROC with good stability and was selected as the feature extractor for POLARIX.

**Notes:**

In addition, we optimized architecture parameters and training hyperparameters. For the fully-connected layer, we compared dimensionalities of 64 and 128 nodes. The 128-node fully-connected layer consistently outperformed the 64-node alternative. Dropout probabilities in both the fully connected network (FCN) and the attention module were varied between 0.25 and 0.50. A dropout rate of 0.50 in both the attention and FCN layers improved generalization compared to lower dropout rates. To address class imbalance in the limited number of *POLE*mut cases, we compared sampling strategies (downsampling, balanced sampling, random sampling) and investigated the use of class-weighted binary cross-entropy loss. Random sampling of training data yielded the highest AUC compared to balanced or downsampling strategies. Use of a class-weighted binary cross-entropy loss further improved results. Finally, we evaluated optimizer parameters, including initial learning rate, learning rate schedule (milestones, gamma), and weight decay. For the optimizer, the optimal configuration was an initial learning rate of 0.0001, decayed by a factor of 0.1 at epochs 5, 15, and 25, with a weight decay of 0.0001. The Adam optimizer was used with default momentum parameters.

The most performant configuration was selected based on mean AUROC across the cross-validation folds. The final model, POLARIX, was trained on a single NVIDIA Quadro RTX 6000 GPU with 24GB of VRAM for 93 epochs in around 19 hours. On average, inference takes 3 minutes for image feature extraction using H-optimus-1[16] and 0.33 seconds for one POLARIX-score prediction using one GPU. The POLARIX model alone (without the H-optimus-1[16] foundation model) contains 984K parameters.

#### 3. Performance experiments

##### 3.1. Robustness checks

*Stain augmentation experiment*

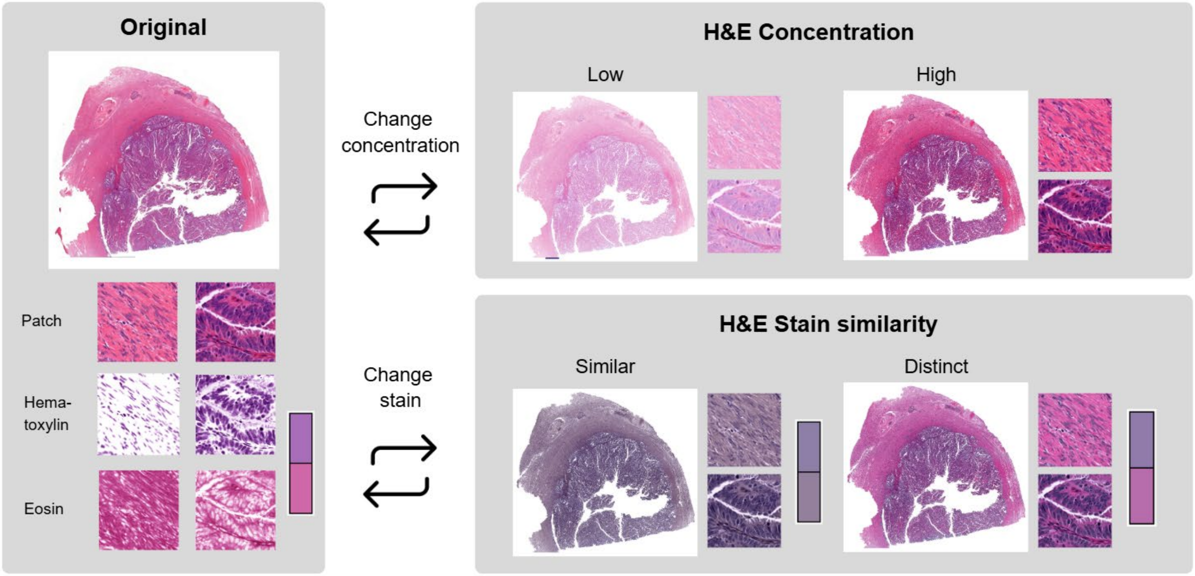


**Figure 3. Methodology of the stain augmentation experiment.** We simulate four corner settings, by augmenting WSIs tile-by-tile to reference H&E concentrations and stains extracted from the Pathology Images of Scanners and Mobilephones (PLISM) dataset[19].

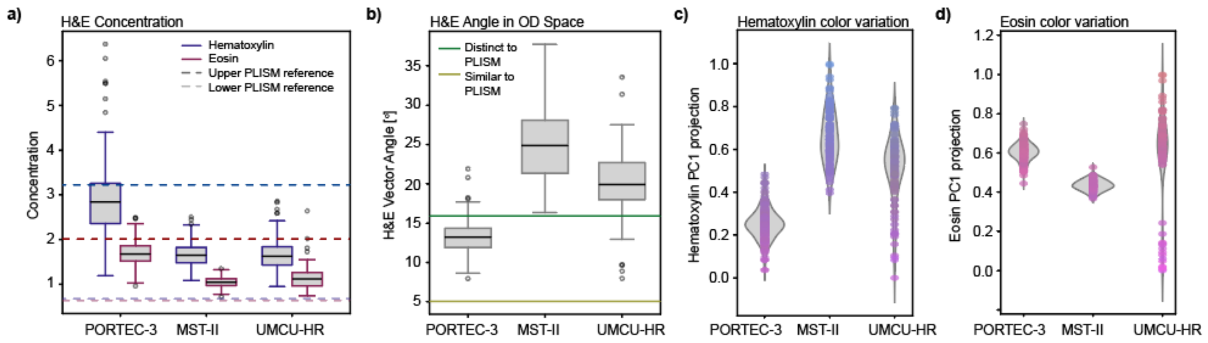


**Figure 4. Inherent cohort stain variation.** a-d) We report the three cohorts variation in concentration (a), H&E vector angle in Optical Density (OD) space (representing the H&E stain similarity, where a lower angle equals a higher similarity) (b), H&E color variation based on the first principal component (PC1) value of the cohorts stain vectors (c-d).

##### *POLARIX-scores and POLE variants*

**Table 4.** POLARIX-score distributions of *POLE* variants in the pooled cohort.

| ***POLE* variant** | ***Pathogeneity*** | ***N.* cases** | **POLARIX-score [mean ± std]** |
|---|---|---|---|
| P286R | pathogenic | 35 | 0.61 ± 0.38 |
| V411L | pathogenic | 15 | 0.50 ± 0.42 |
| A456P | pathogenic | 9 | 0.85 ± 0.30 |
| S297F | pathogenic | 4 | 0.31 ± 0.34 |
| M444K | pathogenic | 1 | 0.99 ± 0 |
| S459F | pathogenic | 3 | 0.60 ± 0.42 |
| G330D | non-pathogenic | 1 | 2.6e-5 |
| D287E | non-pathogenic | 2 | 2.0e-6 ± 1.0e-6 |

**Table 5.** Comparison POLARIX-score distributions between *POLE* variants in the pooled cohort. M444K was not analyzed because it only occurred once.

| **Sample 1** | **Sample 2** | **p-value** | **p-value (Bonferroni)** |
|---|---|---|---|
| A456P | S297F | 0.050 | 0.50 |
| P286R | S297F | 0.12 | 1 |
| P286R | A456P | 0.28 | 1 |
| P286R | V411L | 0.31 | 1 |
| P286R | S459F | 1 | 1 |
| V411L | A456P | 0.14 | 1 |
| V411L | S297F | 0.66 | 1 |
| V411L | S459F | 0.57 | 1 |
| A456P | S459F | 0.73 | 1 |
| S297F | S459F | 0.86 | 1 |

#### 3.2. Fitting the Platt scaler for calibration

**Table 6.** Comparison of calibration metrics before and after applying Platt scaling on the calibration set.

| Method | O:E ratio | Calibration intercept | Calibration slope |
|---|---|---|---|
| POLARIX | 0.82 [0.70,0.94] | -1.02 [-1.54, -0.49] | 0.56 [0.44, 0.68] |
| POLARIX + Platt scaler | 1.14 [0.96, 1.31] | 0.69 [0.11, 1.27] | 0.46 [0.36, 0.56] |

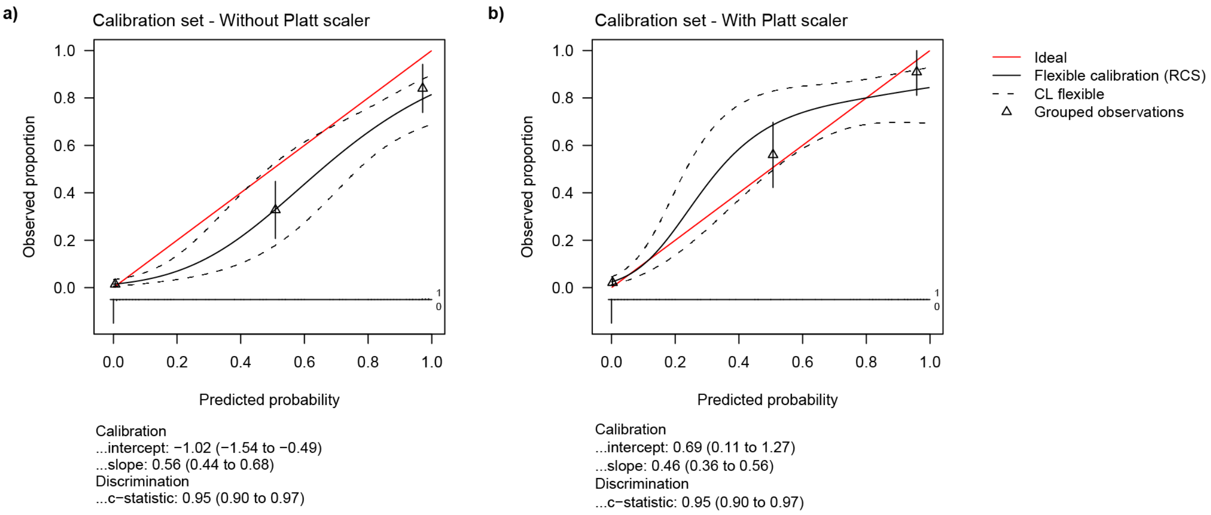


**Figure 5.** Calibration plots with and without Platt Scaling on the calibration hold-out set.

**Notes:**

To assess prediction calibration, we compared POLARIX probabilities in the held-out calibration set with and without Platt scaling[20]. The Platt scaler was fitted on the predictions of the training set of POLARIX (α = 1.21, β= -1.91), and then applied on the hold-out calibration set to prevent data leakage. Calibration performance was evaluated using the observed-to-expected (O:E) ratio, calibration intercept, and calibration slope. In the calibration set, Platt scaling improved overall calibration (**Table 3, Figure 3**). Specifically, the O:E ratio improved from 0.82 (CI: 0.70-0.94) to 1.14 (CI: 0.96-1.31), such that the confidence interval included the perfect 1.00 O:E ratio. The calibration intercept improved from -1.02 (CI: -1.54 - -0.49) to 0.69 (CI: 0.11 – 1.27), i.e., such that it is closer to a perfect intercept result (0). However, the calibration slope did not improve (before Platt scaling: 0.56 (CI: 0.44-0.68), after Platt scaling: 0.46 (CI: 0.36-0.56)). Due to improvements in the O:E ratio and intercept, we chose to use Platt scaling with POLARIX in external testing.

#### 3.3. Calibration assessment

**Table 7.** Calibration metrics of POLARIX on pooled external test cohorts and each test cohort separately.

| Study | Metric | Result |
|---|---|---|
| Pooled | O:E ratio | 1.24 [1.04, 1.49] |
| | Intercept | 1.36 [0.74, 1.97] |
| | Slope | 0.42 [0.33, 0.50] |
| PORTEC-3 | O:E ratio | 1.08 [0.86, 1.32] |
| | Intercept | 0.60 [-0.31, 1.51] |
| | Slope | 0.45 [0.32, 0.58] |
| MST-II | O:E ratio | 2.01 [1.29, 3.27] |

| | | |
|---|---|---|
| | Intercept | 4.17 [2.59, 5.75] |
| | Slope | 0.37 [0.23, 0.52] |
| UMCU-HR | O:E ratio | 1.22 [0.79, 1.83] |
| | Intercept | 0.88 [-0.24, 2.00] |
| | Slope | 0.51 [0.27, 0.75] |

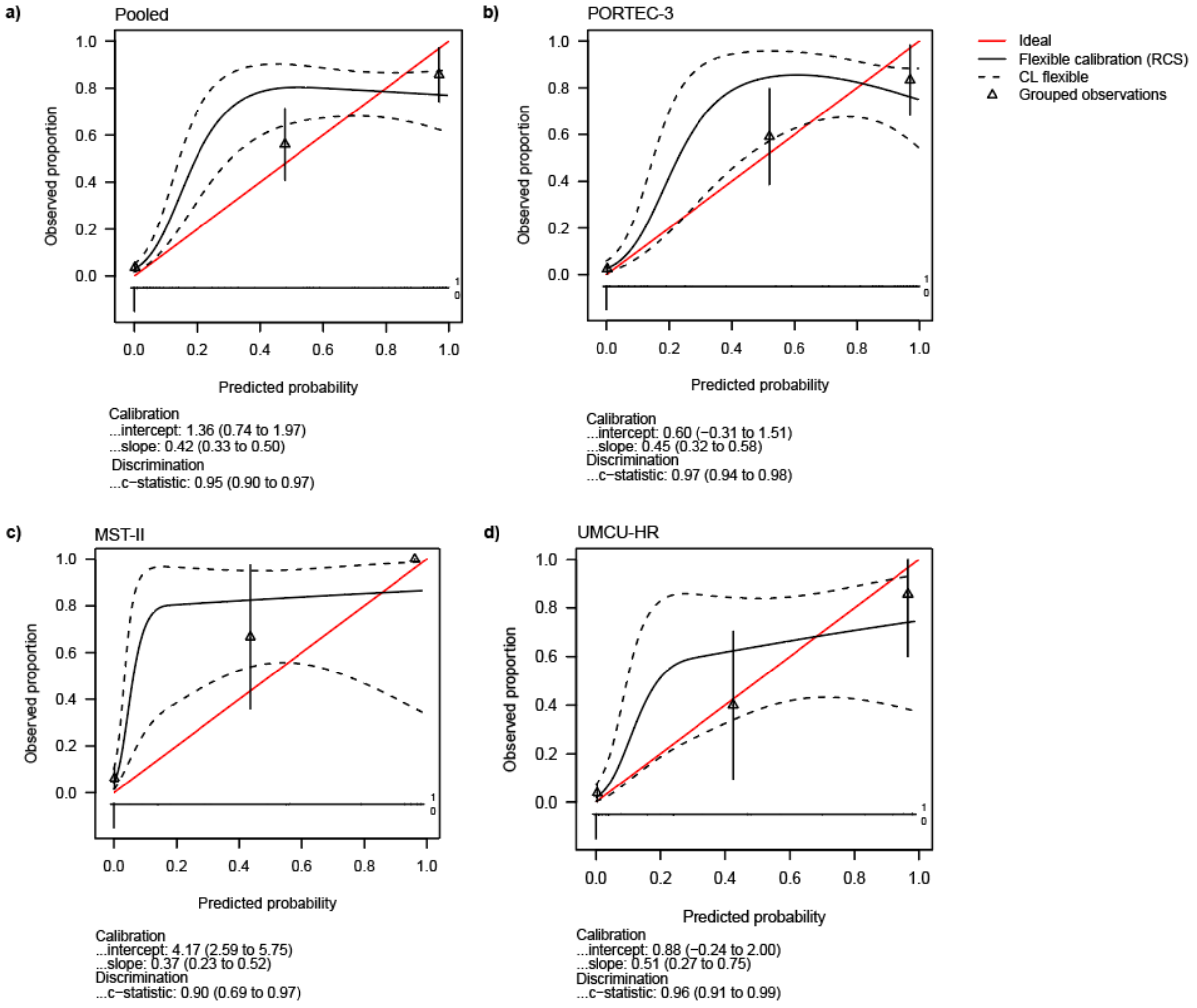


**Figure 6**. Calibration plots of the predictions of all external test cohorts. We found that grouped observations are close to the ideal calibration line. All curves rapidly increase around 0.20, which can be explained by that POLARIX predicts *POLE*wt cases near zero (Pooled: POLARIX-score = 0.014 ± 0.082), and *POLE*mut cases in a higher range (Pooled: POLARIX-score = 0.42 ± 0.38).

#### 3.4. Classification accuracy

**Table 8.** Performance of POLARIX as a screening model. We classified patients into “POLARIX-mut” or “POLARIX-wildtype” groups based on their predicted probabilities and thresholds Low, Mid, and High. Performance criteria were sensitivity, specificity, positive predictive value (PPV), negative predictive value (NPV), and the test reduction in percentages calculated with the true-negative fraction (TN / (TN + FN + FP + TP)). This value approximates how many confirmatory molecular tests could be avoided and should ideally be as high as possible. Conversely, the false-negative fraction (FNF), calculated with FN / (TP + FN) captures the proportion of true *POLE*mut cases missed and should be minimized.

| Test cohort | Threshold (target) | Sensitivity | Specificity | PPV | NPV | Test reduction [%] | FNF |
|---|---|---|---|---|---|---|---|
| Pooled | Low (0.005) | 0.93 [0.85, 0.99] | 0.89 [0.87, 0.92] | 0.59 [0.50, 0.68] | 0.987 [0.973, 0.997] | 77 [73, 81] | 0.07 [0.02, 0.15] |
| | Mid (0.025) | 0.88 [0.80, 0.95] | 0.93 [0.90, 0.95] | 0.66 [0.56, 0.75] | 0.980 [0.966, 0.992] | 80 [76, 83] | 0.12 [0.05, 0.20] |
| | High (0.075) | 0.83 [0.73, 0.91] | 0.94 [0.92, 0.96] | 0.69 [0.59, 0.79] | 0.970 [0.952, 0.985] | 81 [77, 84] | 0.18 [0.09, 0.27] |
| PORTEC-3 | Low (0.005) | 0.97 [0.90, 1.00] | 0.89 [0.85, 0.93] | 0.62 [0.49, 0.74] | 0.995 [0.983, 1.000] | 75 [70, 81] | 0.03 [0.00, 0.09] |
| | Mid (0.025) | 0.92 [0.82, 1.00] | 0.92 [0.88, 0.96] | 0.67 [0.55, 0.80] | 0.985 [0.964, 1.000] | 78 [73, 83] | 0.08 [0.00, 0.18] |
| | High (0.075) | 0.87 [0.75, 0.97] | 0.93 [0.89, 0.96] | 0.67 [0.55, 0.83] | 0.975 [0.950, 0.995] | 79 [74, 84] | 0.13 [0.03, 0.24] |
| MST-II | Low (0.005) | 0.81 [0.60, 1.00] | 0.95 [0.90, 0.99] | 0.72 [0.50, 0.94] | 0.968 [0.926, 1.000] | 81 [74, 88] | 0.19 [0.00, 0.40] |
| | Mid (0.025) | 0.75 [0.50, 0.94] | 0.97 [0.93, 1.00] | 0.80 [0.58, 1.00] | 0.958 [0.916, 0.990] | 83 [76, 89] | 0.26 [0.06, 0.50] |
| | High (0.075) | 0.69 [0.46, 0.91] | 0.97 [0.93, 1.00] | 0.79 [0.56, 1.00] | 0.948 [0.904, 0.989] | 83 [76, 89] | 0.31 [0.08, 0.56] |
| UMCU-HR | Low (0.005) | 0.93 [0.75, 1.00] | 0.85 [0.78, 0.91] | 0.45 [0.27, 0.623] | 0.989 [0.959, 1.000] | 75 [67, 83] | 0.07 [0.00, 0.25] |
| | Mid (0.025) | 0.93 [0.77, 1.00] | 0.90 [0.84, 0.95] | 0.54 [0.33, 0.74] | 0.990 [0.968, 1.000] | 80 [73, 87] | 0.07 [0.00, 0.23] |
| | High (0.075) | 0.86 [0.64, 1.00] | 0.94 [0.89, 0.98] | 0.63 [0.40, 0.84] | 0.981 [0.951, 1.000] | 83 [76, 89] | 0.14 [0.00, 0.36] |

#### 3.5. Net Benefit computation

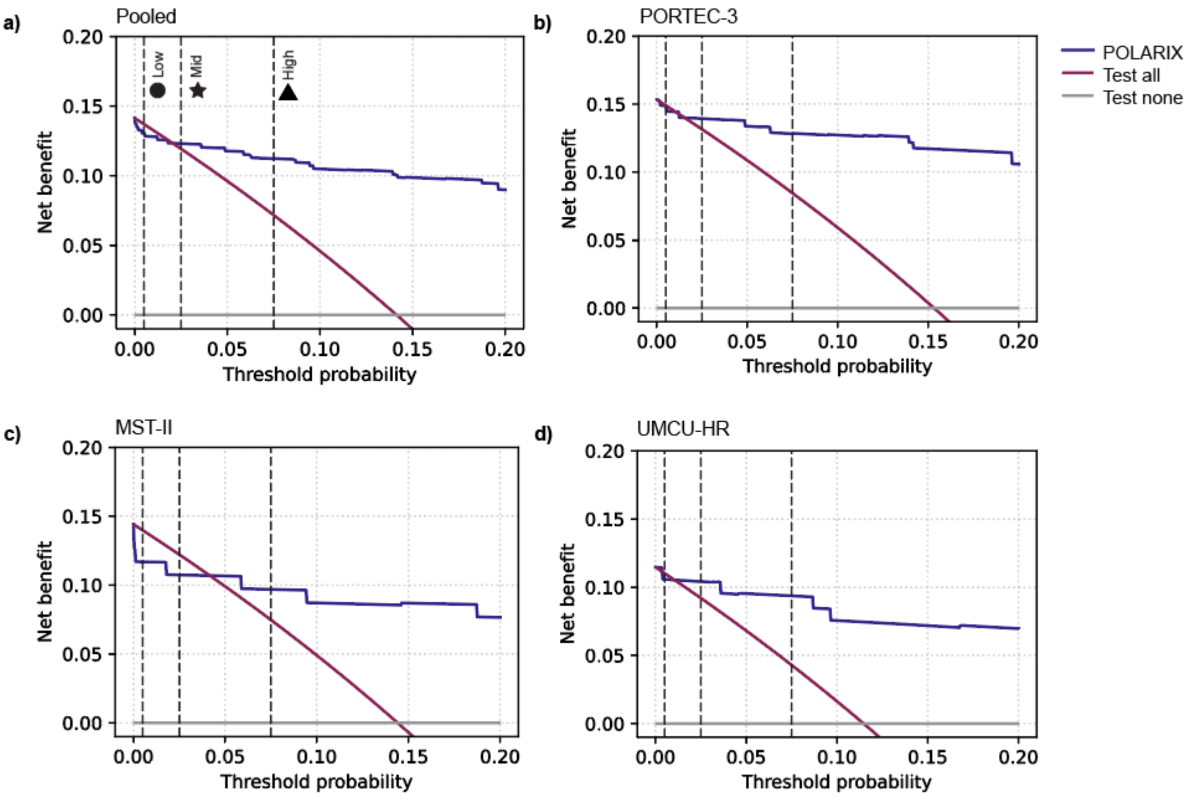


**Figure 7**. Net Benefit plots. Vertical lines depict pre-defined resource-dependent thresholds Low, Mid and High.

**Table 9**. Net Benefit metrics on the three thresholds.

| | | **POLARIX** | | | | **Net Benefit** | | **ΔNet Benefit** | |
|---|---|---|---|---|---|---|---|---|---|
| **Test cohort** | **Threshold** | **TP** | **FP** | **TN** | **FN** | **POLARIX** | **Test All** | **POLARIX vs Test All** | **POLARIX vs Test None** |
| Pooled | Low (0.005) | 63 | 44 | 369 | 5 | 0.131 | 0.137 | -0.007 | **+0.131** |
| | Mid (0.025) | 60 | 31 | 382 | 8 | 0.123 | 0.119 | **+0.004** | **+0.123** |
| | High (0.075) | 56 | 25 | 388 | 12 | 0.112 | 0.072 | **+0.040** | **+0.112** |
| PORTEC-3 | Low (0.005) | 37 | 23 | 187 | 1 | 0.149 | 0.149 | 0.000 | **+0.149** |
| | Mid (0.025) | 35 | 17 | 193 | 3 | 0.139 | 0.132 | **+0.008** | **+0.139** |
| | High (0.075) | 33 | 15 | 195 | 5 | 0.128 | 0.085 | **+0.044** | **+0.128** |
| MST-II | Low (0.005) | 13 | 5 | 90 | 3 | 0.117 | 0.140 | -0.023 | **+0.117** |
| | Mid (0.025) | 12 | 3 | 92 | 4 | 0.107 | 0.122 | -0.015 | **+0.107** |
| | High (0.075) | 11 | 3 | 92 | 5 | 0.097 | 0.075 | **+0.022** | **+0.097** |
| UMCU-HR | Low (0.005) | 13 | 16 | 92 | 1 | 0.106 | 0.110 | -0.004 | **+0.106** |
| | Mid (0.025) | 13 | 11 | 97 | 1 | 0.104 | 0.092 | **+0.012** | **+0.104** |
| | High (0.075) | 12 | 7 | 101 | 2 | 0.094 | 0.043 | **+0.051** | **+0.094** |

### 4. Explainability experiments

#### 4.1. Clinicopathological correlates of POLARIX-scores

**Table 10.** Multiple linear regression was performed to evaluate the association between clinicopathological variables and the continuous POLARIX score. Categorical variables were coded using Stage I, Grade 1, and EEC as reference groups (coefficients represent adjusted mean differences relative to these groups).

| **Variable** | | Coefficient (95% CI) | p-value |
|---|---|---|---|
| Intercept | | 0.20 (0.08, 0.33) | 0.0011 |
| p53 IHC | Mutant | -0.26 (-0.37, -0.15) | 3.5e-06 |
| Stage | I | 1 (reference) | |
| | II | -0.13 (-0.25, -0.02) | 0.020 |
| | III | -0.15 (-0.25, -0.05) | 0.0047 |
| Grade | GR1 | 1 (reference) | |
| | GR2 | 0.03 (-0.08, 0.15) | 0.57 |
| | GR3 | 0.30 (0.17, 0.42) | 3.8e-06 |
| Histotype | Endometrioid | 1 (reference) | |
| | Clear cell carcinoma | -0.24 (-0.43, -0.04) | 0.019 |
| | Serous | -0.09 (-0.20, 0.027) | 0.13 |
| | Other | -0.041 (-0.26, 0.18) | 0.71 |
| LVSI | Present | -0.015 (-0.091, 0.061) | 0.70 |
| L1CAM | Positive | -0.062 (-0.18, 0.055) | 0.30 |
| ER | Negative | -0.006 (-0.11, 0.096) | 0.91 |
| PR | Negative | 0.044 (-0.055, 0.14) | 0.38 |

#### 4.2. Attention analyses

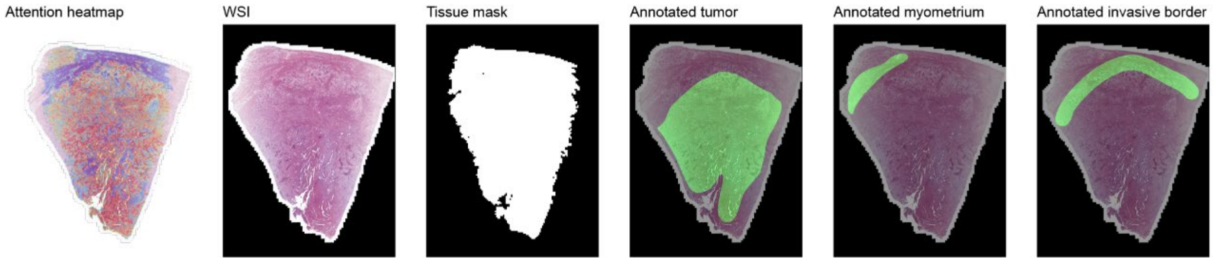


**Figure 8.** Example of a WSI with its attention heatmap, tissue mask, and annotated regions. Heatmaps of WSIs were generated by first segmenting the tissue and extracting non-overlapping tiles. Each tile was represented by its H-optimus-1 embedding and fed into POLARIX, yielding one attention score per patch. Attention scores were standardized at the slide level and mapped onto a lower-resolution image using the Jet colormap from OpenCV, having red for high attention and blue for low attention. To produce visually continuous heatmaps, the resulting score maps were smoothed with a Gaussian filter before overlaying on the slide image. Highly attended tiles in slides were extracted using the coordinates of the tiles with the highest raw attention scores. In total, 393 WSIs in cohort PORTEC-3 were annotated by an expert GYN-pathologist (TB) using QuPath(16) version 0.2.0 (17) for three regions: the tumor region, invasive border and myometrium[21]. The normalized attention per region was computed by taking the sum of the attention scores in the region of interest and dividing by its size in pixels. In this way, this score was not dependent on the size of the region, limiting human-introduced bias, and we can compare the attention across the regions.

#### 4.3. CellViT++ deployment and feature analysis

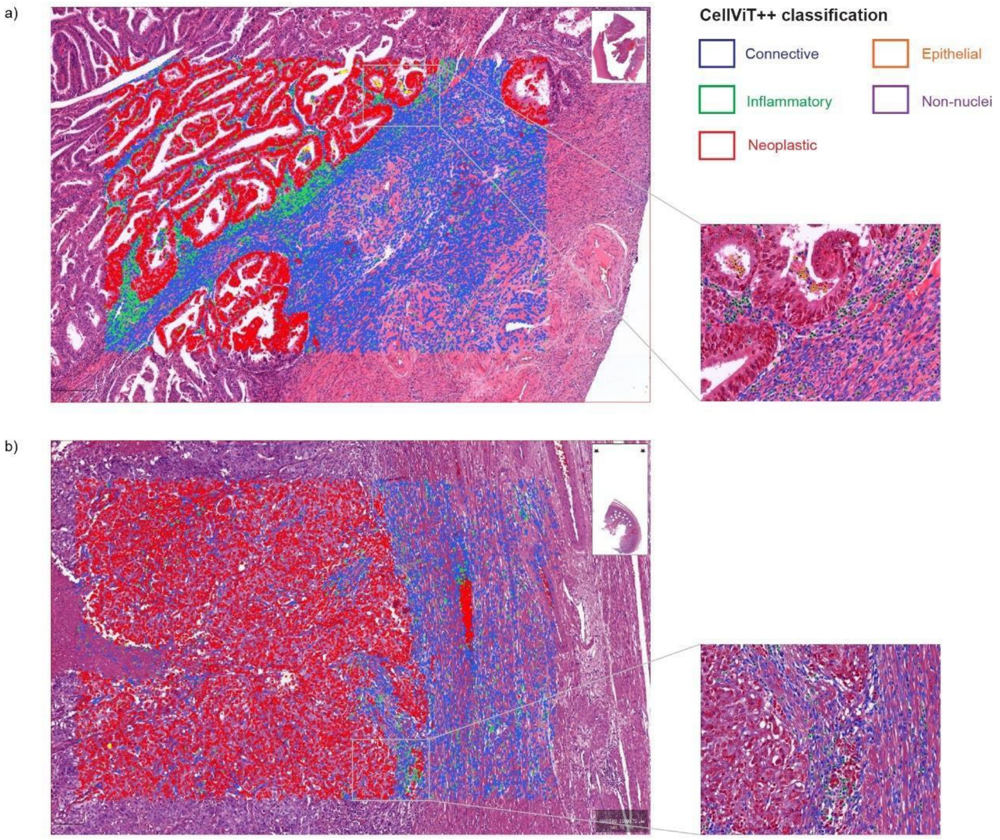


**Figure 9.** CellViT++ classification results are visualized with an overlay of the classifications over the WSI in two cases (a and b) with zoomed-in regions. To count the number of inflammatory cells and to compute the nuclear atypia score, we used CellViT++ to classify and segment detected cells in WSIs. We used the PanNuke classification, which includes connective, epithelial, inflammatory, non-nuclei and neoplastic cells. We did not finetune the network, because the non-finetuned model already yielded satisfactory classification accuracy. All cells on a WSI were classified and for each cell, metrics including nuclei area size, sphericity and convexity were calculated. These results were aggregated onto slide-level per cell type using the median and IQR.

##### *Nuclear Atypia Score*

We define the nuclear atypia score as the average of non-convexity and non-sphericity, which we computed for the neoplastic (tumor) cells as follows:

$$Nuclear\ Atypia\ Score\ =\ \frac{1}{2} *\ [(1-convexity)\ +\ (1-sphericity)],$$

where convexity is calculated as

$$convexity\ =\ Area_{object}\ /\ Area_{convex\ hull}$$

and sphericity is calculated as

$$sphericity\ =\ 2 * \sqrt{\pi * Area_{object}}\ /\ Perimeter.$$

#### 4.4. Misclassification Review in PORTEC-3

**Table 11.** Comparison of distributions of prediction groups of clinicopathological covariates found significantly correlated with POLARIX-scores (Fisher 2x2 exact test). Prop. TN = proportion of true negatives, prop. FP = proportion of false positives, prop. FN = proportion of false negatives, prop. TP = proportion of true positives.

| **Variable** | **Threshold** | **prop. TN** | **prop. FP** | **prop. FN** | **prop. TP** | **TP vs FP (p-value)** | **TP vs TN (p-value)** | **FP vs TN (p-value)** | **FN vs TP (p-value)** | **FN vs TN (p-value)** |
|---|---|---|---|---|---|---|---|---|---|---|
| p53 IHC mutant | Low | 81/187 (0.43) | 9/23 (0.39) | 0/1 (0.00) | 6/37 (0.16) | 0.067 | 0.0017 | 0.82 | 1.0 | 1.0 |
| | Mid | 83/193 (0.43) | 7/17 (0.41) | 0/3 (0.00) | 6/35 (0.17) | 0.089 | 0.0043 | 1.0 | 1.0 | 0.26 |
| | High | 84/195 (0.43) | 6/15 (0.40) | 1/5 (0.20) | 5/33 (0.15) | 0.075 | 0.0019 | 1.0 | 1.0 | 0.40 |
| FIGO Stage II | Low | 55/187 (0.29) | 7/23 (0.30) | 1/1 (1.00) | 5/37 (0.14) | 0.1830 | 0.066 | 1.0 | 0.16 | 0.30 |
| | Mid | 57/193 (0.30) | 5/17 (0.29) | 1/3 (0.33) | 5/35 (0.14) | 0.2644 | 0.066 | 1.0 | 0.41 | 1.0 |
| | High | 58/195 (0.30) | 4/15 (0.27) | 1/5 (0.20) | 5/33 (0.15) | 0.43 | 0.095 | 1.0 | 1.0 | 1.0 |
| FIGO Stage III | Low | 85/187 (0.45) | 11/23 (0.48) | 0/1 (0.00) | 9/37 (0.24) | 0.091 | 0.018 | 0.83 | 1.0 | 1.0 |
| | Mid | 89/193 (0.46) | 7/17 (0.41) | 1/3 (0.33) | 8/35 (0.23) | 0.20 | 0.015 | 0.80 | 1.0 | 1.0 |
| | High | 90/195 (0.46) | 6/15 (0.40) | 1/5 (0.20) | 8/33 (0.24) | 0.32 | 0.022 | 0.79 | 1.0 | 0.38 |
| CCC | Low | 9/187 (0.05) | 0/23 (0.00) | 0/1 (0.00) | 1/37 (0.03) | 1.0 | 1.0 | 0.60 | 1.0 | 1.0 |
| | Mid | 9/193 (0.05) | 0/17 (0.00) | 0/3 (0.00) | 1/35 (0.03) | 1.0 | 1.0 | 1.0 | 1.0 | 1.0 |

| | | | | | | | | | | |
|---|---|---|---|---|---|---|---|---|---|---|
| | High | 9/195 (0.05) | 0/15 (0.00) | 0/5 (0.00) | 1/33 (0.03) | 1.0 | 1.0 | 1.0 | 1.0 | 1.0 |
| GR3 | Low | 98/187 (0.52) | 12/23 (0.52) | 0/1 (0.00) | 34/37 (0.92) | 0.0010 | 0.0000 | 1.0 | 0.11 | 0.48 |
| | Mid | 101/193 (0.52) | 9/17 (0.53) | 2/3 (0.67) | 32/35 (0.91) | 0.0029 | 0.0000 | 1.0 | 0.29 | 1.0 |
| | High | 103/195 (0.53) | 7/15 (0.47) | 4/5 (0.80) | 30/33 (0.91) | 0.0017 | 0.0000 | 0.79 | 0.45 | 0.37 |

**Table 12.** Comparison of distributions of prediction groups of morphological variables significantly correlated with POLARIX-scores.

| Variable | Threshold | n. TN | mean +- SD TN | n. FP | mean +- SD FP | n. FN | mean +- SD FN | n. TP | mean +- SD TP | TP vs FP | TP vs TN | FP vs TN | FN vs TP | FN vs TN |
|---|---|---|---|---|---|---|---|---|---|---|---|---|---|---|
| Inflammatory Fraction | Low | 187 | 0.08 ± 0.05 | 23 | 0.10 ± 0.06 | 1 | 0.05 ± nan | 37 | 0.10 ± 0.05 | 0.9636 | 0.0009 | 0.0313 | 0.2105 | 0.5638 |
| | Mid | 193 | 0.08 ± 0.05 | 17 | 0.09 ± 0.05 | 3 | 0.08 ± 0.06 | 35 | 0.10 ± 0.05 | 0.7550 | 0.0020 | 0.1090 | 0.4680 | 0.8166 |
| | High | 195 | 0.08 ± 0.05 | 15 | 0.09 ± 0.05 | 5 | 0.07 ± 0.04 | 33 | 0.11 ± 0.05 | 0.6404 | 0.0011 | 0.1557 | 0.1226 | 0.9335 |
| CD8+ T Cell Density | Low | 132 | 5.14 ± 1.77 | 11 | 7.02 ± 1.77 | 0 | NA | 25 | 7.98 ± 2.00 | 0.1492 | 0.0000 | 0.0018 | NA | NA |
| | Mid | 133 | 5.15 ± 1.77 | 10 | 7.02 ± 1.87 | 0 | NA | 25 | 7.98 ± 2.00 | 0.1709 | 0.0000 | 0.0040 | NA | NA |
| | High | 133 | 5.15 ± 1.77 | 10 | 7.02 ± 1.87 | 1 | 8.78 ± nan | 24 | 7.94 ± 2.03 | 0.1922 | 0.0000 | 0.0040 | 0.8800 | 0.0448 |
| TLS Count | Low | 180 | 0.21 ± 0.84 | 23 | 0.22 ± 0.67 | 1 | 0.00 ± nan | 34 | 1.74 ± 2.94 | 0.0019 | 0.0000 | 0.4842 | 0.3687 | 0.7793 |

| | | | | | | | | | | | | | | |
|---|---|---|---|---|---|---|---|---|---|---|---|---|---|---|
| | Mid | 186 | 0.20 ± 0.83 | 17 | 0.29 ± 0.77 | 2 | 0.00 ± 0.00 | 33 | 1.79 ± 2.97 | 0.0103 | 0.0000 | 0.2030 | 0.1841 | 0.6868 |
| | High | 188 | 0.20 ± 0.83 | 15 | 0.33 ± 0.82 | 3 | 0.33 ± 0.58 | 32 | 1.81 ± 3.01 | 0.0244 | 0.0000 | 0.1298 | 0.3615 | 0.1421 |
| Median Nuclei Size | Low | 187 | 30.00 ± 6.40 | 23 | 25.99 ± 3.71 | 1 | 28.99 ± nan | 37 | 27.03 ± 3.81 | 0.3267 | 0.0035 | 0.0009 | 0.4117 | 0.9706 |
| | Mid | 193 | 29.83 ± 6.41 | 17 | 26.52 ± 3.61 | 3 | 29.31 ± 1.07 | 35 | 26.89 ± 3.87 | 0.7698 | 0.0042 | 0.0145 | 0.1164 | 0.8899 |
| | High | 195 | 29.79 ± 6.39 | 15 | 26.68 ± 3.83 | 5 | 29.83 ± 1.91 | 33 | 26.67 ± 3.83 | 0.9291 | 0.0025 | 0.0358 | 0.0221 | 0.6527 |
| Nuclear Atypia Score | Low | 187 | 0.07 ± 0.01 | 23 | 0.07 ± 0.01 | 1 | 0.06 ± nan | 37 | 0.07 ± 0.00 | 0.0127 | 0.0000 | 0.2083 | 0.6842 | 0.1915 |
| | Mid | 193 | 0.07 ± 0.01 | 17 | 0.07 ± 0.01 | 3 | 0.06 ± 0.00 | 35 | 0.07 ± 0.00 | 0.1233 | 0.0000 | 0.0811 | 0.1086 | 0.0026 |
| | High | 195 | 0.07 ± 0.01 | 15 | 0.07 ± 0.01 | 5 | 0.06 ± 0.00 | 33 | 0.07 ± 0.00 | 0.2958 | 0.0000 | 0.0896 | 0.0212 | 0.0001 |
| Mitotic Density | Low | 186 | 22.51 ± 14.05 | 23 | 26.04 ± 16.33 | 1 | 15.00 ± nan | 37 | 27.86 ± 13.33 | 0.5082 | 0.0050 | 0.2914 | 0.2540 | 0.6564 |
| | Mid | 192 | 22.35 ± 14.13 | 17 | 29.12 ± 15.41 | 3 | 18.33 ± 7.57 | 35 | 28.31 ± 13.46 | 0.9844 | 0.0033 | 0.0447 | 0.1363 | 0.8608 |
| | High | 194 | 22.54 ± 14.18 | 15 | 27.53 ± 15.76 | 5 | 20.40 ± 6.84 | 33 | 28.61 ± 13.78 | 0.5856 | 0.0048 | 0.1559 | 0.1667 | 0.8564 |

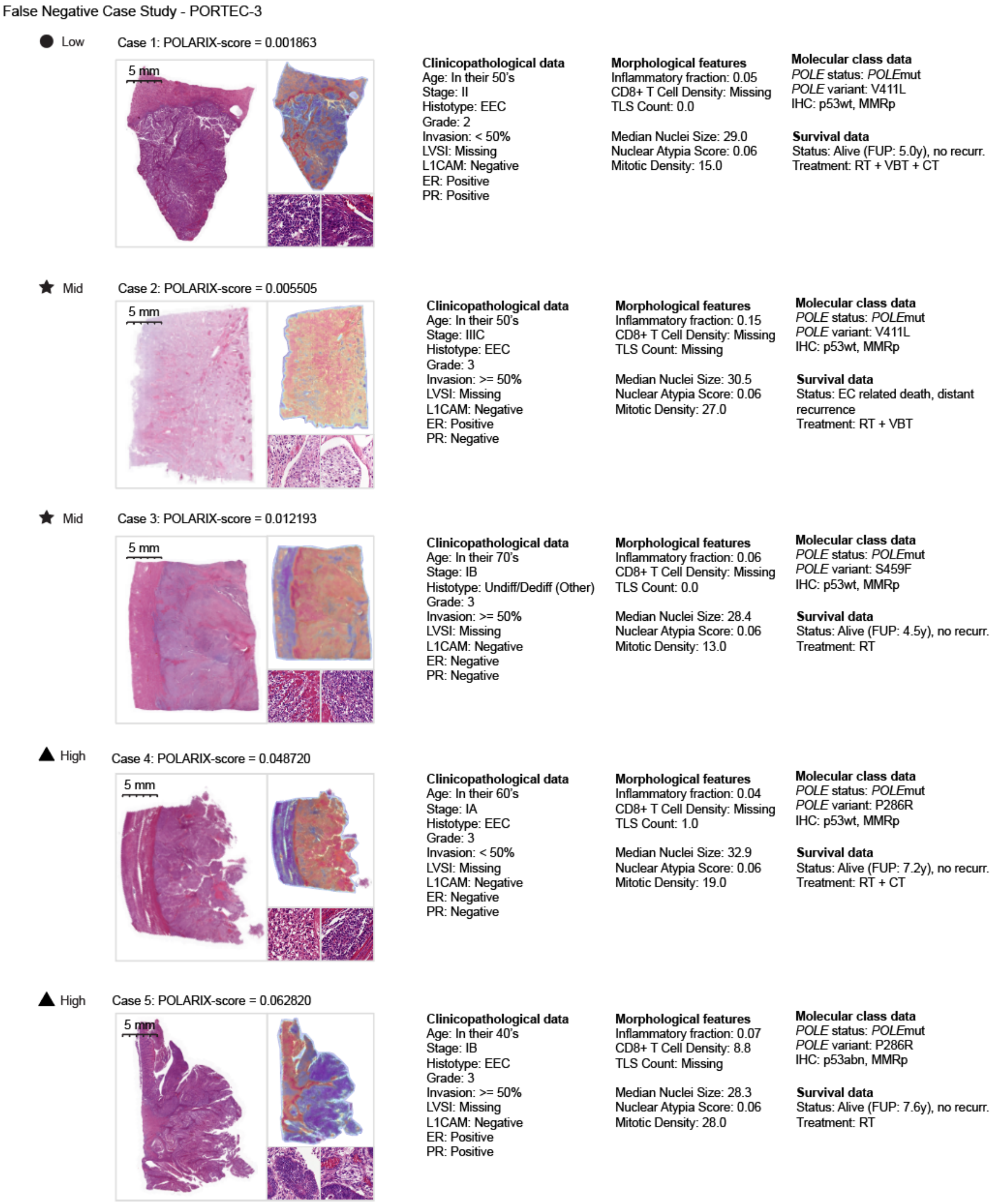


**Figure 10.** False Negative case study in PORTEC-3 showing the WSI, heatmap, clinicopathological data, morphological features, molecular class data, and survival data.

#### 4.5. Clinical outcome experiments

**Table 13**. Five-year overall recurrence-free estimates of the simulation of implementation of POLARIX in PORTEC-3.

| Test cohort | Threshold | S(t) *POLE*mut [%] (95%CI) | S(t) *POLE*wt [%] (95%CI) | S(t) POLARIX-wt [%] (95%CI) | Logrank p-value |
|---|---|---|---|---|---|
| PORTEC-3 | Low (0.005) | 97 (82-100) | 70 (47-84) | 65 (58-72) | $2.6 \times 10^{-5}$ |
| | Mid (0.025) | 100 (100-100) | 71 (43-87) | 65 (58-71) | $6.2 \times 10^{-5}$ |
| | High (0.075) | 100 (100-100) | 73 (44-89) | 65 (58-72) | $1.7 \times 10^{-4}$ |

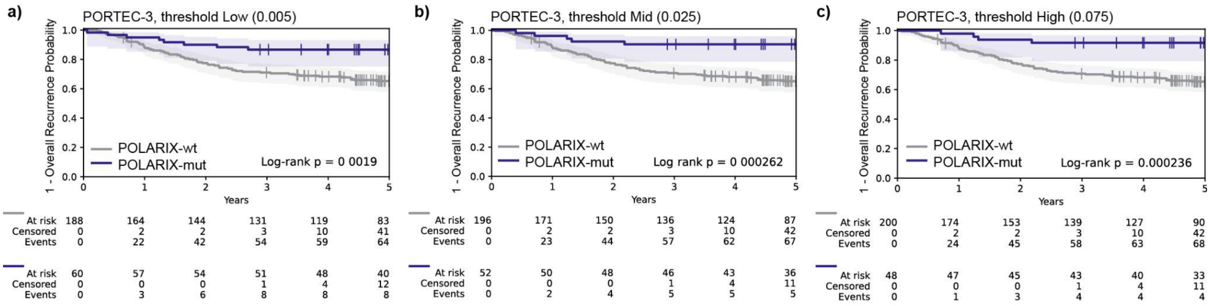


**Figure 11.** Kaplan-Meier analysis on PORTEC 3 (a-c) on thresholds Low, Mid and High stratified by POLARIX-mut (blue) and POLARIX-wt (grey). The log-rank test was used to compute differences in clinical outcomes.

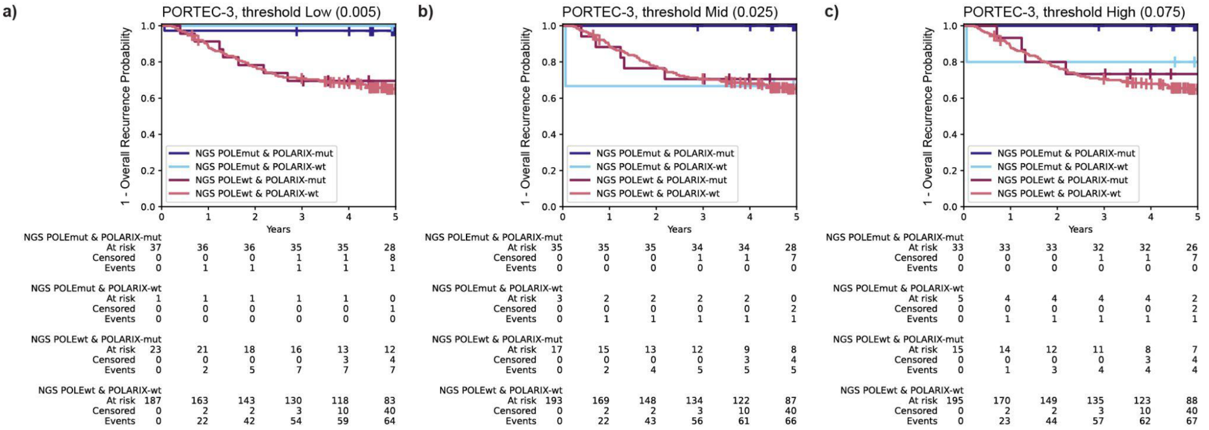


**Figure 12.** Kaplan-Meier analysis done on PORTEC 3 (a-c) on thresholds Low, Mid and High stratified by NGS result and POLARIX prediction groups.

### 5. Sample size calculation

We performed a sample size calculation using the precision method. The standard error of the AUC was approximated using the method described by Hanley and McNeil[22], using the observed *POLE*mut to *POLE*wt ratio (r = 68/481 = 0.165). Assuming a true AUC of 0.95 and targeting a 95% CI ≥ 0.90, this approach indicated that at least 37 POLEmut cases and 221 POLEwt cases (total n = 257) were required.

In detail, this was:

$$SE_{target} = \frac{(AUC - 95\%CI_{lower\ bound})}{1.96} = \frac{(0.95 - 0.90)}{1.96} = 0.0255$$

$$r = \frac{n_1}{n_0} = \frac{68}{481} = 0.165,$$

where $n_1$= number of cases diseased (POLEmut) and $n_0$= number of control cases (POLEwt)

By using the target SE as the *SE(AUC),* the assumed true AUC and ratio of control and diseased cases, we solved the following equation:

$$SE(AUC) = \sqrt{\frac{AUC(1 - AUC) + (n_1 - 1)(Q_1 - AUC^2) + (n_0 - 1)(Q_2 - AUC^2)}{n_0 n_1}},$$

where

$$Q_1 = \frac{AUC}{2 - AUC} \text{ and } Q_2 = \frac{2AUC^2}{1 + AUC}.$$

This yielded a minimum of 221 control cases and 37 event cases.
